# *Wolbachia*-mediated sterility suppresses *Aedes aegypti* populations in the urban tropics

**DOI:** 10.1101/2021.06.16.21257922

**Authors:** The Project Wolbachia – Singapore Consortium, Ng Lee Ching

## Abstract

Incompatible insect technique (IIT) via releases of male *Wolbachia*-infected mosquitoes is a promising tool for dengue control. In a three-year trial in Singaporean high-rise housing estates, we demonstrated that *Wolbachia-*based IIT dramatically reduces both wildtype *Aedes aegypti* populations [reductions of 92.7% (95% CI: 84.7%–95.8%) and 98.3% (97.7%–99.8%)] and dengue incidence [reductions of 71% (43%-87%) to 88% (57%-99%)] in the targeted areas. The study highlights the need to ensure adequate vertical distribution of released males in high-rise buildings, address immigration of wildtype females from neighboring areas, and prevent and mitigate stable establishment of *Wolbachia* in field mosquito populations. Our results demonstrate the potential of *Wolbachia-*based IIT (supplemented with irradiation, in Singapore’s context) for strengthening dengue control in tropical cities, where dengue burden is the greatest.

**One Sentence Summary:** Releases of male *Wolbachia-*infected *Aedes aegypti* suppress dengue mosquitoes and reduce dengue incidence in high-rise urban areas in Singapore.

## Main Text

Incompatible insect technique (IIT) (*1*), a promising complementary strategy for the control of arbovirus transmission, involves the release of male mosquitoes infected with *Wolbachia, a* maternally inherited endosymbiotic bacterium. Due to cytoplasmic incompatibility (CI) (*2, 3*), matings between *Wolbachia*-infected males and wildtype females not infected with the same *Wolbachia* strain yield non-viable eggs. Repeated releases of *Wolbachia*-infected males are thus expected to suppress wildtype mosquito populations and reduce disease transmission. Like classical sterile insect technique (SIT), where releases of irradiation-sterilized males have led to large-scale elimination of agricultural pests (*4, 5*), IIT avoids disadvantages associated with traditional vector control, including genetic or behavioral resistance to insecticides, off-target effects, and the inability to locate cryptic larval sites. IIT further avoids fitness costs arising from exposure to male-sterilizing doses of irradiation, which may reduce the mating competitiveness of released sterile males in an SIT program (*6*).

However, IIT approaches require measures to minimize unintentional release (due to imperfect sex-sorting) of fertile *Wolbachia-*infected female mosquitoes, which may lead to stable establishment of the released *Wolbachia* strain in field mosquito populations (*7, 8*). While *Wolbachia-*established populations pose no public health or ecological threat, and in fact confer a public health benefit due to their reduced ability to transmit dengue (*9, 10*), establishment renders CI-based IIT ineffective, and may result in increased biting pressure if the *Wolbachia-*established population is not well controlled.

To reduce the likelihood of stable establishment, two recent pilot trials combined IIT with SIT, using low-dose irradiation to sterilize residual females during releases of *Wolbachia-*infected males; this IIT-SIT approach suppressed *Aedes aegypti* populations in semi-rural village settings in Thailand (*11*), and *Aedes albopictus* populations in riverine islands in Guangzhou, China (*12*). Another successful pilot study in low-rise residential neighborhoods in California, US, employed a sex-sorting pipeline that uses industrial vision software and machine learning to drastically reduce the probability of female contamination and stable establishment (*13*).

Compared to the low-rise, relatively isolated, or semi-rural study sites in the abovementioned pilots (*11*–*13*), the highly urbanized city-state of Singapore presents a more challenging landscape for IIT and dengue control in general. Singapore is one of the most densely populated nations in the world, with an average of 7,866 people per square kilometer (*14*) and about 95 percent of households living in high-rise apartments (*15*). This landscape, combined with the tropical climate, is ideal for year-round reproduction of *Aedes aegypti*, the primary dengue vector (*16*). With 19.1 million visitor arrivals in 2019 (*17*), Singapore’s position as a travel hub facilitates the introduction and circulation of new dengue virus strains as well as other emerging *Aedes-*borne viruses such as Zika, some of which have high epidemic potential (*18, 19*). Further, decades of effective integrated vector control focused on source reduction (*20, 21*) have lowered population immunity against dengue (*22*), leaving the country vulnerable to explosive dengue outbreaks (*23, 24*).

To augment Singapore’s existing vector control program with new tools, we conducted an extensive field trial of IIT targeting *Aedes aegypti*. While Singapore’s low *Aedes aegypti* population (*21*) is advantageous for further suppression by IIT, it also increases the likelihood of stable establishment of *Wolbachia* in field *Aedes aegypti* (*25, 26*). In our trial, we thus used either IIT-SIT or a high-fidelity sex-sorting methodology (*13*) to minimize the likelihood of stable establishment. Releases of male *Wolbachia-*infected mosquitoes suppressed *Aedes aegypti* populations and reduced dengue incidence in both our field sites, which are located in high-rise housing estates. The results demonstrate the feasibility of IIT for use in a tropical urban context.

### A local *Wolbachia-*infected *Aedes aegypti* line, *w*AlbB-SG, for IIT

We generated *w*AlbB-SG, a localized *Aedes aegypti* line stably infected with the *w*AlbB strain of *Wolbachia*, by outcrossing SG (a wildtype *Aedes aegypti* line derived from field collections in Singapore) with WB2 (an *Aedes aegypti* line of Texas, US origin stably trans-infected with *w*AlbB) for six generations.

We observed 100% maternal transmission and 100% CI induced by *w*AlbB in *w*AlbB-SG. All 1,648 offspring from 103 female *w*AlbB-SG screened were *w*AlbB*-*positive. Although *w*AlbB density was significantly higher in younger males (Fig. S1A), all eggs produced from crosses between wildtype SG females and 3-, 6-, and 11-day-old male *w*AlbB-SG were non-viable (Fig. S1B). The *w*AlbB-SG and SG lines showed similar response profiles to pyrethroid and organophosphate insecticides (Fig. S1C). In addition, *w*AlbB-SG males demonstrated equivalent mating performance to SG in cage assays, with a competitiveness index (C) (*27*) of 1.1. Taken together, these results demonstrate the suitability of *w*AlbB-SG as an IIT line for the control of *Aedes aegypti* populations in Singapore.

### Phase 1: Understanding the behavior of *w*AlbB-SG in the field

Following approval from Singapore’s Ministry of Sustainability and the Environment and extensive community engagement (*28*), we conducted small-scale releases in historically dengue high-risk sites (*29*) in Yishun and Tampines high-rise housing estates (Fig. 1A) to characterize the bionomics of *w*AlbB-SG males in Singapore’s urban environment. For this Phase, control sites were located 260m and 1,750m away from the Yishun and Tampines treatment sites respectively. Size-based sex-sorting was performed at the pupae stage using the Fay-Morlan glass plate separation method (*30*), with a female contamination rate of ∼1 in 1000.

**Figure 1.**
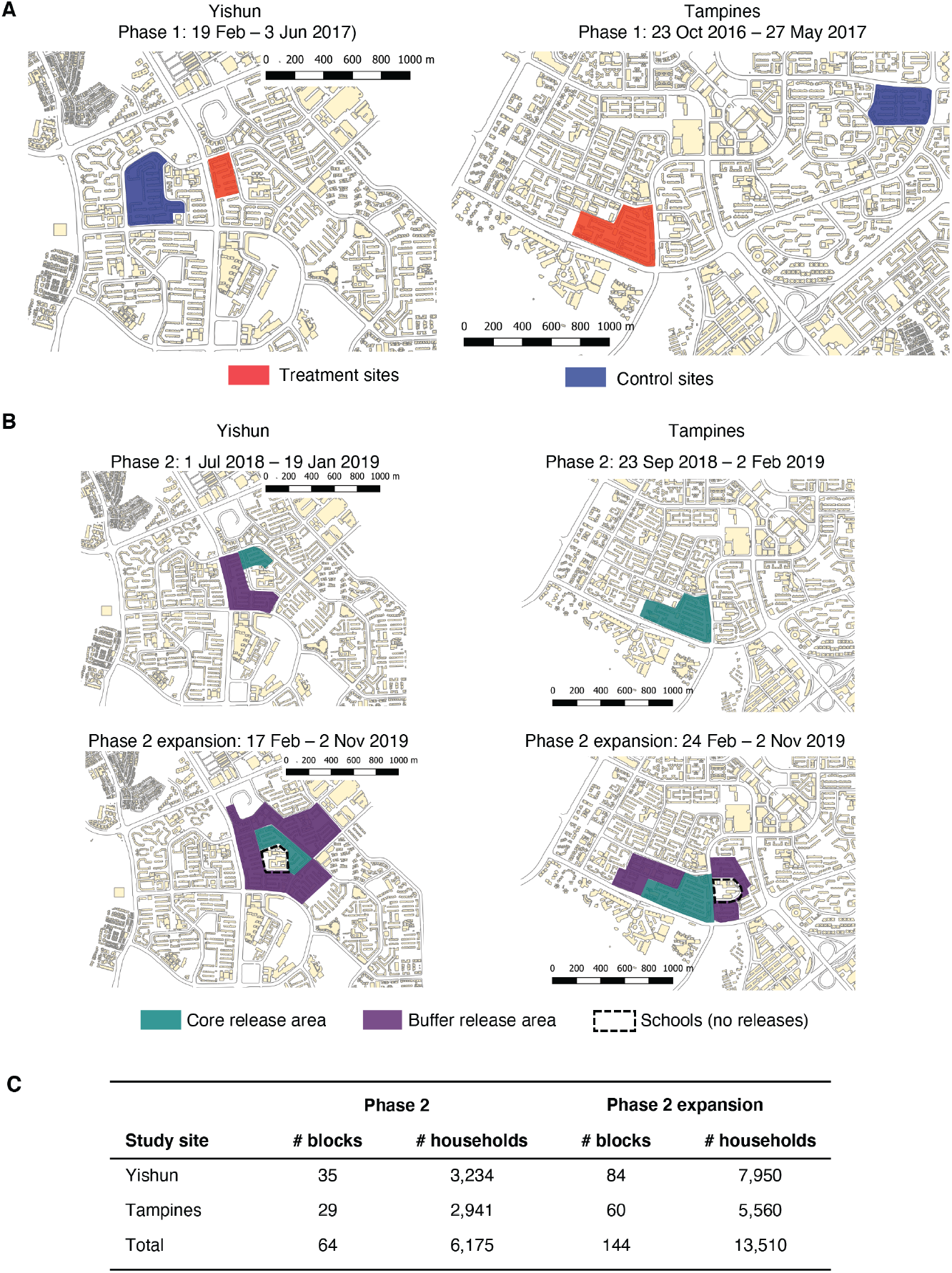
Study sites for field releases of *w*AlbB-SG males. **(A)** Map of treatment and control sites in the Phase 1 small-scale study in Yishun and Tampines high-rise housing estates. **(B)** Map of core and buffer treatment zones in the Phase 2 suppression trial. Treatment areas were expanded over the course of the trial. **(C)** Numbers of high-rise apartment blocks and households covered by releases in the Phase 2 suppression trial.

Weekly ground-floor releases were carried out for 15 weeks in Yishun and 31 weeks in Tampines. During the releases, we observed a significant (p<0.001) 91% and 66% reduction in the hatch rates of eggs collected in the Yishun and Tampines release sites respectively, as compared to control sites (Fig. 2A, C). Hatch rates in the release and control sites were not significantly different during the pre- and post-intervention periods (Fig. 2A, C), indicating that they bounced back quickly once releases stopped. Moreover, the adult female *Aedes aegypti* population, estimated by the Gravitrap *Aedes aegypti* Index (GAI, the average number of *Aedes aegypti* females caught per Gravitrap (*31*)) was suppressed by up to 51% and 65% in the Yishun and Tampines release sites compared to control sites (Fig. 2B, D), as measured by before-after control-impact (BACI) analysis. This limited suppression can be attributed to the poor vertical distribution of released *w*AlbB-SG males, which was skewed towards lower floors, in contrast to the more evenly distributed wildtype females (Fig. S2A). Other reasons may include the short average lifespan of the males (∼4.5 days), which would result in a reduced *w*AlbB-SG field population for a few days prior to the next weekly release, as well as immigration of wildtype mosquitoes from contiguous non-release areas. Adult female numbers also bounced back quickly after releases stopped (Fig. 2B, D). The quick rebound of hatch rates and adult mosquito population post-release could be due to residual local population and/or immigration of wildtype females and males facilitated by the small size of the sites.

**Figure 2.**
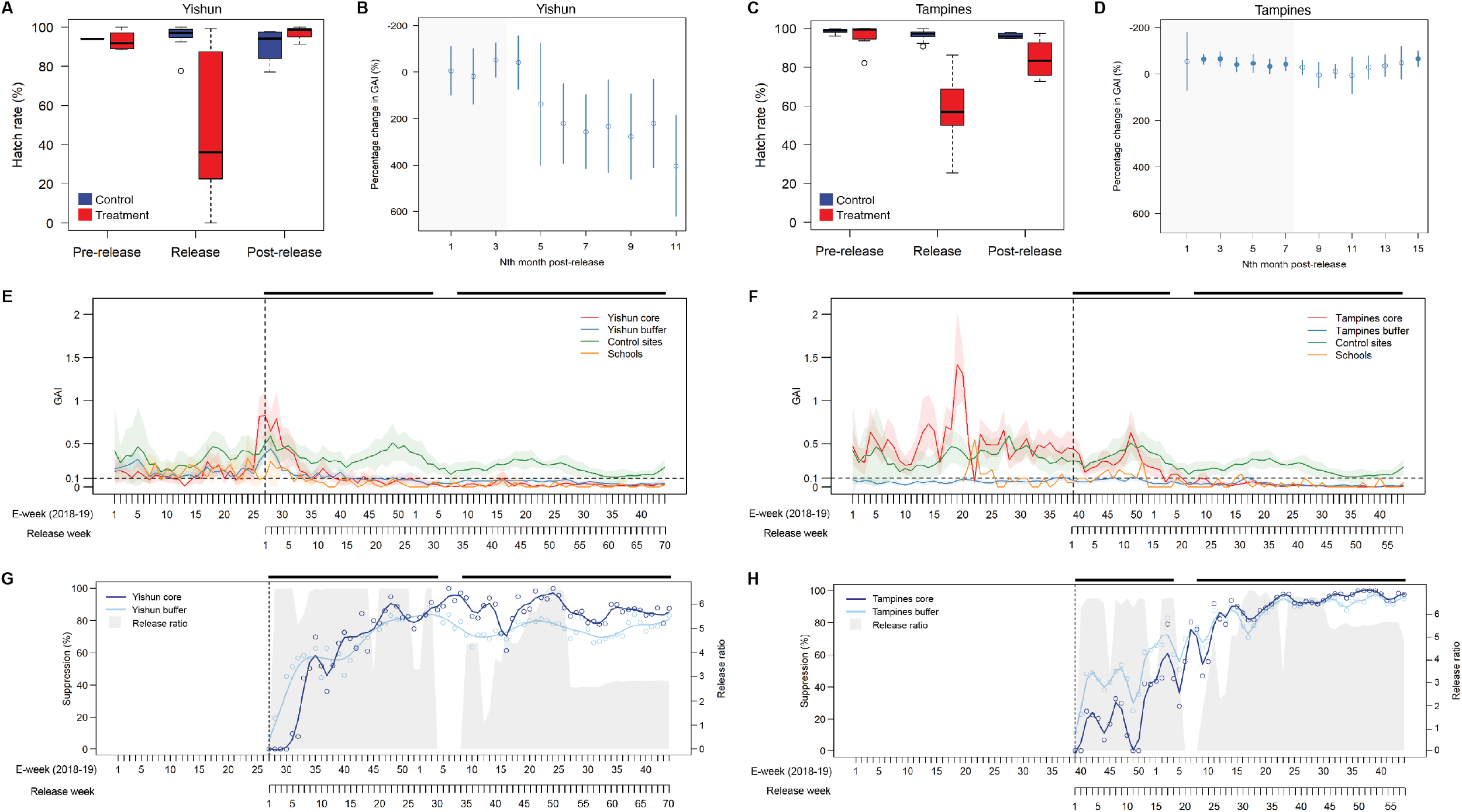
Suppression of *Aedes aegypti* mosquito populations following Phase 1 (A-D) and Phase 2 (E-H) releases of *w*AlbB-SG males. **(A)** Weekly egg hatch rates in the Yishun treatment and control sites pre-, during, and post-Phase 1 releases. Lines and limits of the boxes indicate medians and 25^th^ and 75^th^ percentiles respectively; whiskers extend to 1.5 times the interquartile range. Outliers are shown as points. **(B)** Monthly BACI suppression estimates of GAI in the Yishun treatment site during (grey-shaded region) and post-Phase 1 releases. Point estimates and 95% confidence intervals are shown; negative values indicate suppression. Filled circles indicate statistical significance (p<0.05). **(C-D)** Same as (A-B) but for the Tampines treatment and control sites. **(E-F)** Weekly GAI in control sites and in core release zones, buffer release zones, and schools in the Yishun **(E)** and Tampines **(F)** Phase 2 study sites. Schools did not receive releases. Shaded areas indicate 95% CIs. The horizontal dotted line indicates the low-dengue risk GAI threshold of 0.1. **(G-H)** Weekly adult female *Aedes aegypti* suppression compared to control sites and weekly release ratios (number of *w*AlbB-SG males released per resident per week) in the Yishun **(G)** and Tampines **(H)** Phase 2 study sites. The start of Phase 2 releases is indicated by the vertical dotted line, and release periods are denoted by horizontal black bars. Release sites were expanded following the pause in releases.

Towards the final four weeks of the release period, *w*AlbB-positive *Aedes aegypti* larvae were regularly detected in ovitraps in the treatment sites. This continued for 8 and 11 weeks post-release in Yishun and Tampines respectively, averaging 2.2% of ovitraps per week. Similarly, we detected a small number of *w*AlbB-positive female adults up to 7 and 14 weeks post-release in Yishun and Tampines, respectively. These results demonstrate the potential for *Wolbachia* establishment in field populations due to imperfect sex-sorting. No *w*AlbB-positive larvae were detected in the subsequent six weeks of ovitrap deployment, and no *w*AlbB-positive female adults were detected in the subsequent 38 weeks, suggesting that *w*AlbB mosquitoes failed to establish in the field as the wildtype population bounced back. This agrees with a previous study that showed that a rather high threshold of ∼20% *w*AlbB in the field population was needed to effect *w*AlbB invasion and establishment (*8, 32, 33*).

### Phase 2 suppression trial: mitigating imperfect sorting and ecological challenges

Phase 2, conducted 13 and 16 months after the final Phase 1 release in Yishun and Tampines respectively, aimed to suppress *Aedes aegypti* in expanded sites (Fig. 1B) through mitigating the likelihood of *w*AlbB establishment, addressing the short lifespan and poor vertical distribution of released males, and counteracting immigration of wildtype mosquitoes. In Yishun, we adopted IIT-SIT (*11, 12*), treating Fay-Morlan-sorted *w*AlbB-SG male pupae with low-dose X-ray irradiation to render residual females sterile (100% female sterility was achieved at 30 Gray, with 0 out of 92 mated females laying eggs). Irradiation had no impact on *w*AlbB density in *w*AlbB-SG males (Fig. S1D), and we continued to observe 100% CI induction after irradiation (Fig. S1E-F). Laboratory studies showed that irradiated *w*AlbB-SG males and their non-irradiated counterparts had similar average lifespans (∼7 days) (Fig. S1G), but irradiated *w*AlbB-SG males had reduced mating competitiveness (Fig. S1F).

In Tampines, as a second approach to mitigate *w*AlbB establishment, we used a high-fidelity sex-sorting pipeline to reduce the female contamination rate to ∼1×10^−9^, six orders of magnitude lower than Fay-Morlan sorting (*13*). Pipeline-sorted *w*AlbB-SG males displayed comparable life expectancy and mating competitiveness with Fay-Morlan-sorted males (Fig. S1H-I) and higher mating competitiveness compared to irradiated males (Fig. S1F, I).

Males were released along the ground floor and an upper floor (level 5 and upwards) of each block, yielding a more even vertical dispersal of the mosquitoes than with ground-floor releases alone (Fig. S2). Two releases a week were performed, which modelling suggested would maintain steadier field populations of *w*AlbB-SG males in treatment sites as compared to weekly releases (Fig. S3). Release areas roughly doubled in size over the course of the Phase 2 as core release areas were surrounded with buffer releases (buffer areas) to counteract immigration of females from contiguous non-release areas (Fig. 1B-C); in some cases, existing geographical features such as four-lane roads served as barriers.

Seven “sentinel” sites with comparable historical dengue risk and significantly correlated baseline GAI (Fig. S4) served as controls; these sites are distributed across Singapore and similarly located in high-rise public housing estates (*34*).

### Suppression of urban *Aedes aegypti* populations by male *w*AlbB-SG

In high-rise housing estates in Singapore, a GAI<0.1 is associated with significantly lower odds of dengue transmission (*35*). GAI in the Yishun and Tampines cores dropped from >0.8 and >0.4 at the start of releases to below the 0.1 low-risk threshold by release week 9 and 17 respectively (Fig. 2E-F). From this point, aside from minor fluctuations, GAI in both cores was maintained at <0.1 until the end of Phase 2 (release weeks 10–70 for Yishun and 18–58 for Tampines). These sustained periods of low mosquito population had not been observed in four and two years of pre-intervention baseline data for Yishun and Tampines respectively. Correspondingly, high suppression levels (80–100%) compared to control sites were sustained for multiple consecutive weeks in both cores (Fig. 2G-H).

Sustained low mosquito populations were also seen in buffer areas. GAI in the Yishun buffer fell from >0.4 to <0.1 by release week 17, and was maintained at ∼0.1 or below until the end of Phase 2 (Fig. 2E). The low starting GAI in the Tampines buffer (∼0.1 or below) was maintained throughout Phase 2 (Fig. 2F). By comparison, GAI in the control sites fluctuated between 0.1– 0.6, never falling below the 0.1 threshold (Fig. 2E-F).

At the start of Phase 2, approximately 6–7 *w*AlbB-SG males were released per study site resident per week (Fig. 2G-H), corresponding to an initial estimated overflooding ratio of 30:1 *w*AlbB-SG males to wildtype males. Despite a pause in releases and a temporary dip in release numbers in early 2019, no rebound in the core GAIs was observed and suppression levels remained high (Fig. 2G-H), perhaps due to the enlargement of the site to include buffer releases by this point (Fig. 1B-C), and because the overflooding ratio remained high as wildtype male populations decreased due to suppression. Likewise, the Yishun core and buffer GAIs remained <0.1 and high suppression was maintained (Fig. 2E, G) even after release numbers were intentionally halved from July 2019 onwards (Fig. 2G). This suggests that, once achieved over a sufficiently large and well-buffered area, suppression can be maintained more efficiently (i.e. with lower-volume releases), even when using an IIT-SIT strain with reduced fitness (Fig. S1F).

The robust suppression observed was further supported by BACI analysis, which adjusts for baseline differences between treatment and control areas, seasonal effects, and secular trends (Fig. 3). BACI analysis showed a statistically significant 80% suppression of GAI in the Yishun core by the fourth month post-intervention; by the end of Phase 2 (17 months), suppression was sustained at >90% (Fig. 3A). In Tampines, statistically significant suppression (41%) was seen in the first month post-intervention; suppression was sustained at >90% from month 8 to the end of Phase 2 (month 14) (Fig. 3C). These suppression estimates were robust in additional sensitivity analyses (Table S1). We observed a 92.7% reduction in GAI (95% CI 84.7%–95.8%) over Phase 2 in the Yishun core, and a 98.3% reduction in GAI (95% CI 97.7%–99.8%) over Phase 2 in the Tampines core. In contrast, the counterfactual scenarios suggested that the GAI would remain mostly >0.1 in the absence of any release (Fig. 3B, D).

**Figure 3.**
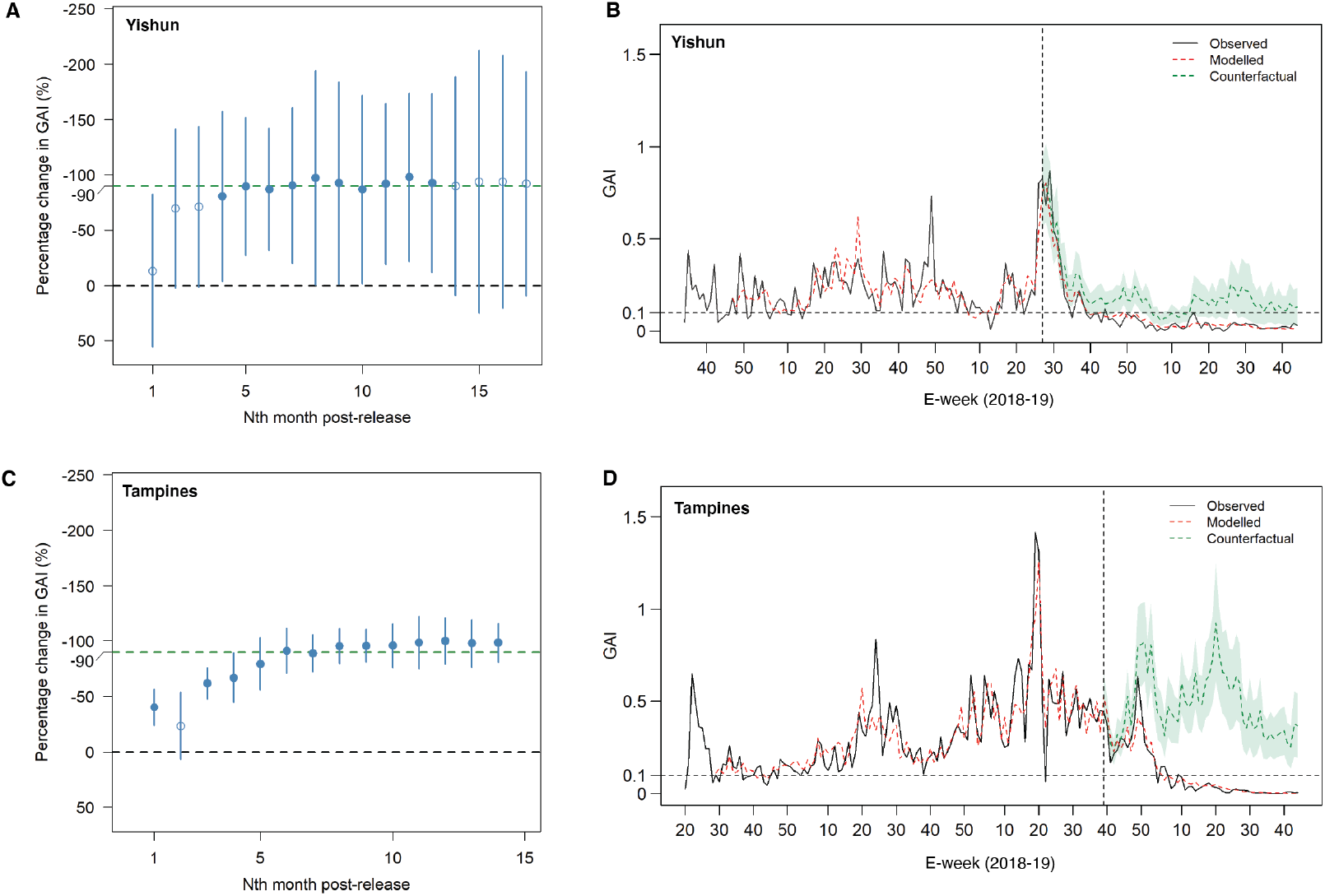
BACI analysis of adult female *Aedes aegypti* suppression. **(A, C)** Monthly BACI suppression estimates of GAI in the Yishun **(A)** and Tampines **(C)** cores over Phase 2. Point estimates and 95% confidence intervals are shown; negative values indicate suppression. Filled circles indicate statistical significance (p<0.05). **(B, D)** Observed, modelled, and counterfactual GAI trends in the Yishun **(B)** and Tampines **(D)** cores pre- and post-Phase 2. The modelled and counterfactual trendlines were produced using a negative binomial regression model. Shaded areas indicate 95% CIs; the horizontal and vertical dotted lines indicate the low-dengue risk GAI threshold of 0.1 and the start of Phase 2 releases, respectively.

### Reduction in dengue incidence within and beyond release areas

In addition to suppressed *Aedes aegypti* populations, core and buffer release zones in Yishun and Tampines showed 71% (95% CI 43%-87%) to 88% (95% CI 57%-99%) lower dengue incidences in 2019 compared to control sites (p<0.001), while no significant differences in dengue incidence between release and control areas were seen pre-trial in 2017 (Fig. 4A-B, Tables S2, S3). Core and buffer release zones also showed lower 2019 dengue incidences compared to non-release areas within the same town (statistically significant for Yishun, p=0.039 and <0.001 for core and buffer areas respectively) (Table S2). These results are consistent with the prolonged maintenance of GAI in release areas below the low-dengue risk threshold of 0.1.

**Figure 4.**
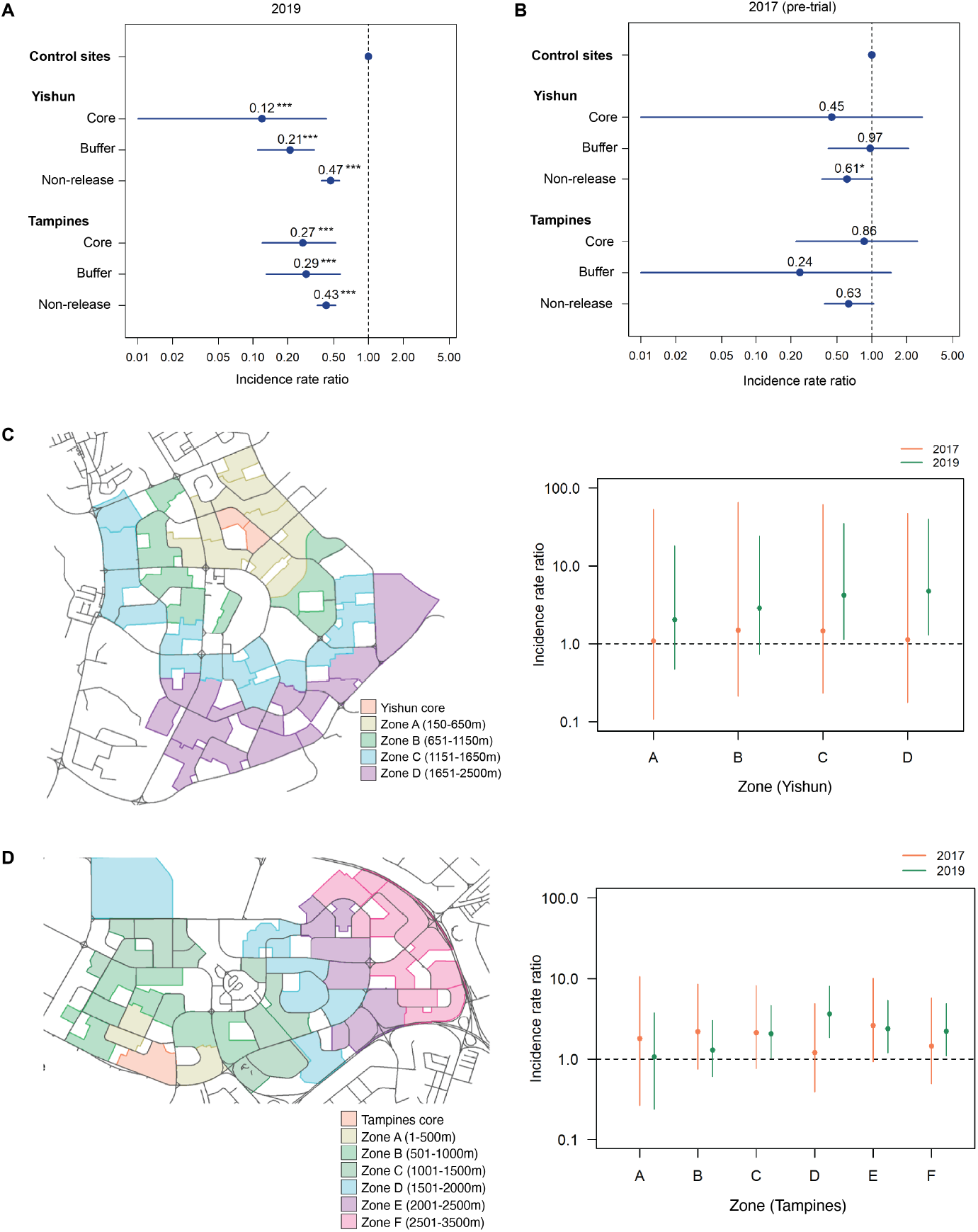
Reduction in dengue incidence at Phase 2 release sites and potential spillover impact on non-release areas. **(A-B)** Rate ratios of dengue incidence in core release zones, buffer release zones, and non-release areas in Yishun and Tampines towns, compared to control sites in 2019 **(A)** and 2017 **(B)**. Point estimates and 95% confidence intervals are shown. ***, p<0.001; *, p<0.05. **(C-D)** Rate ratios of dengue incidence in non-release zones relative to core release zones in Yishun **(C)** and Tampines **(D)**. Point estimates and 95% confidence intervals are shown.

We further observed that even non-release areas in Yishun and Tampines showed significantly lower 2019 dengue incidences than control sites (Fig. 4A, Table S2). It is possible that Yishun and Tampines coincidentally experienced lower dengue transmission compared to control sites in 2019. However, we also observed that both dengue incidence and GAI in the Yishun and Tampines non-release areas increased with distance away from the cores, a trend that was absent in pre-trial dengue incidence data (pre-trial GAI data in non-release areas was not available) (Fig. 4C-D, Fig. S5). This implies that the lower dengue incidence observed in non-release areas may in part be attributed to positive spillover effects from neighboring release areas. Our data suggests that this spillover extended up to ∼1km away from the cores (Fig. 4C-D, Fig. S5).

### Suppression of mosquito populations due to spillover effect of releases

Schools located within the sites (Fig. 1B) did not directly receive releases of *w*AlbB-SG males but were completely surrounded by release areas. Similar to the cores, GAI in the schools (which covered an area of approximately 4.39 hectares in Yishun and 3.40 hectares in Tampines) fell and was maintained at ∼0.1 or below for most of Phase 2 (Fig. 2E-F). This could be attributed to the buffering effect of surrounding releases, which may have counteracted female immigration, and to a positive spillover effect caused by immigration of *w*AlbB-SG males from surrounding releases into the schools.

Similar positive spillover effect can also be seen elsewhere in the study sites (Fig. 5). In Yishun, upon the start of releases in zones 4, 5, and 7, suppression in the adjacent zones 1-3, 6, and 8 was seen even before these zones received releases (Fig. 5A). Similarly in Tampines, partial coverage of zones 1, 4, and 5 was sufficient for prolonged 80-100% suppression in these zones (Fig. 5B). The spillover effect is supported by data from Phase 1 indicating that about 19% of recovered *w*AlbB-SG males were caught outside release sites, up to 230m from release points.

**Figure 5.**
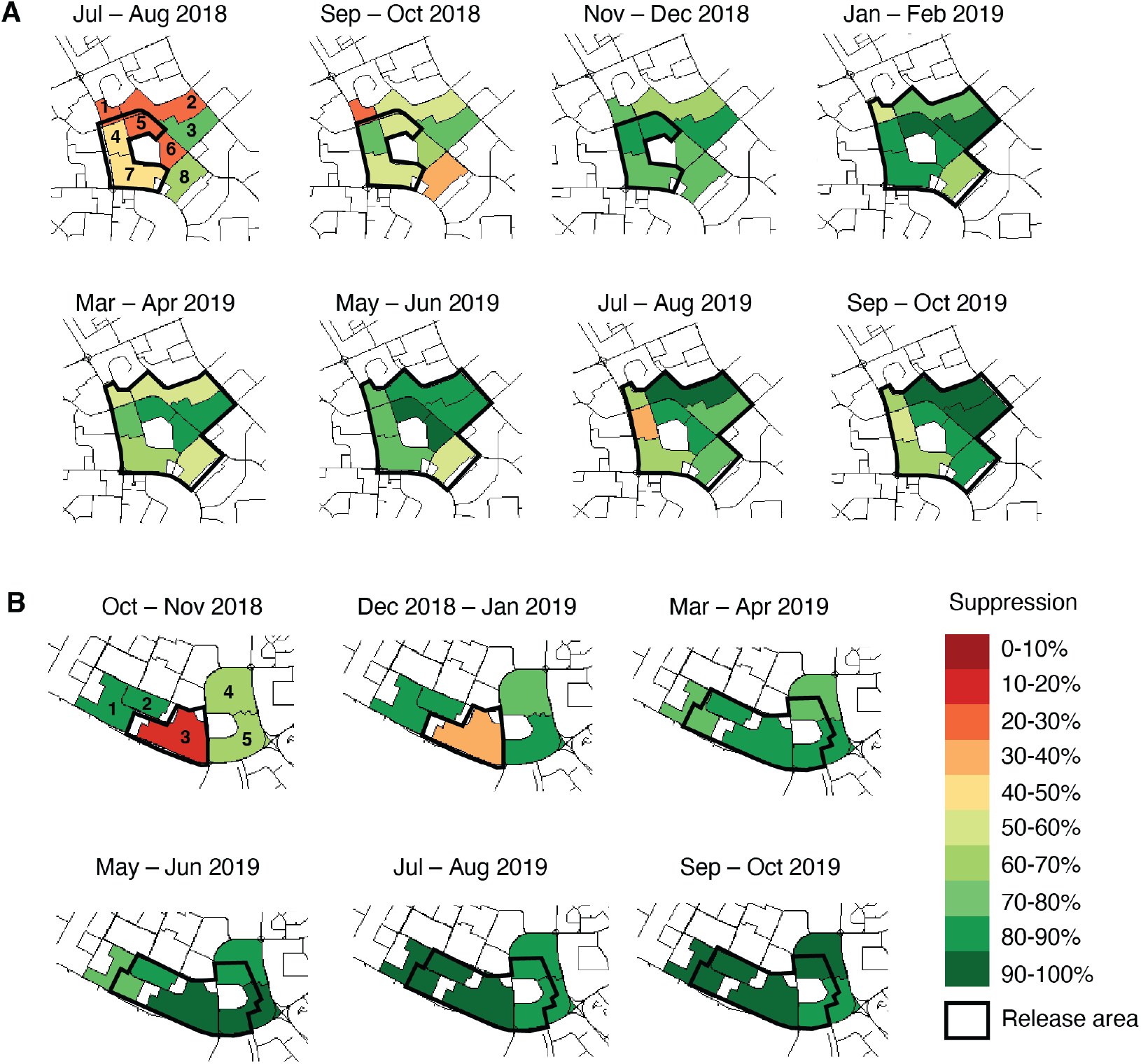
Spatial variation in adult female *Aedes aegypti* suppression. GAI suppression levels compared to control sites in various zones in the Yishun **(A)** and Tampines **(B)** study sites.

Despite high overall suppression, we observed spatial variation in *Aedes aegypti* suppression within the release sites (Fig. 5). Zones located in the interior of release sites (zones 5 and 6 in Yishun; zone 3 in Tampines, i.e. the cores) showed consistently high suppression. Despite being bordered by a major road, zones 1, 4, and 7 in Yishun; and zones 4 and 5 in Tampines had lower and less consistent suppression levels. Areas adjacent to high-footfall areas (zones 7 and 8 in Yishun, located just north of a large mall and bus interchange) had similar inconsistent suppression levels (Fig. 5). This suggests that the relatively small roads in Singapore may not act as migration barriers and the risk of mosquito immigration may be higher in high-traffic areas, making them more resistant to suppression.

### Wildtype *Aedes aegypti* population resistant to elimination

We did not observe elimination of the wildtype *Aedes aegypti* population in the core areas of release sites, suggesting that suppression was less effective in high-traffic buffer areas (Fig. 2, 5) and that immigration of wildtype females from surrounding non-release areas was occurring. Based on estimated suppression levels over the entire release area at the end of Phase 2 (55.9% for Yishun and 91.2% for Tampines, as determined by BACI analysis), we developed a compartmental model (Fig. S6) for female immigration. The model estimated that the daily average number of females migrating to the release area from the surrounding non-release area was roughly 22.7% and 2.8% of the pre-release total number of females in the release area for Yishun and Tampines respectively, assuming that immigration was the sole factor that led to the observed number of mosquitoes remaining in the field. In our model, the suppression effect does not increase linearly with the number of mosquitoes being released, assuming near-constant total immigration from non-release areas. Once the ratio of *w*AlbB-SG males to wildtype males reaches a threshold of ∼8, increasing the size of release has a limited but additional impact on suppression levels. The weaker overall suppression observed in Yishun was therefore unlikely to have been due to the reduction in release numbers at this site (Fig. 2G). Rather than increasing release numbers, achieving better suppression may require the effects of immigration to be negated by further expanding the release site to target additional non-release areas. Community vigilance against mosquito breeding should also enhance suppression.

### X-ray irradiation prevents and mitigates *w*AlbB establishment

After Phase 2, we gradually expanded releases in the Yishun and Tampines sites to include adjacent areas (Fig. S7). We continued to monitor mosquito populations and dengue incidences in the Yishun and Tampines cores. As releases of irradiated IIT-SIT *w*AlbB-SG males in Yishun had a 0.1–0.3% female contamination rate, we consistently detected low levels of *w*AlbB-positive adult *Aedes aegypti* females in the Yishun core during Phase 2 and beyond. However, the use of irradiation to sterilize residual females prior to release prevented stable establishment. This is supported by only rare findings of *w*AlbB-positive larvae in the field and continued suppression in the Yishun core for more than two years (Fig. 6A). Along with the sustained low mosquito population, dengue incidence in the Yishun core was also reduced by 90% in 2020 compared to control sites (incidence rate ratio 0.10, 95% CI 0.02-0.31, p<0.001).

**Figure 6.**
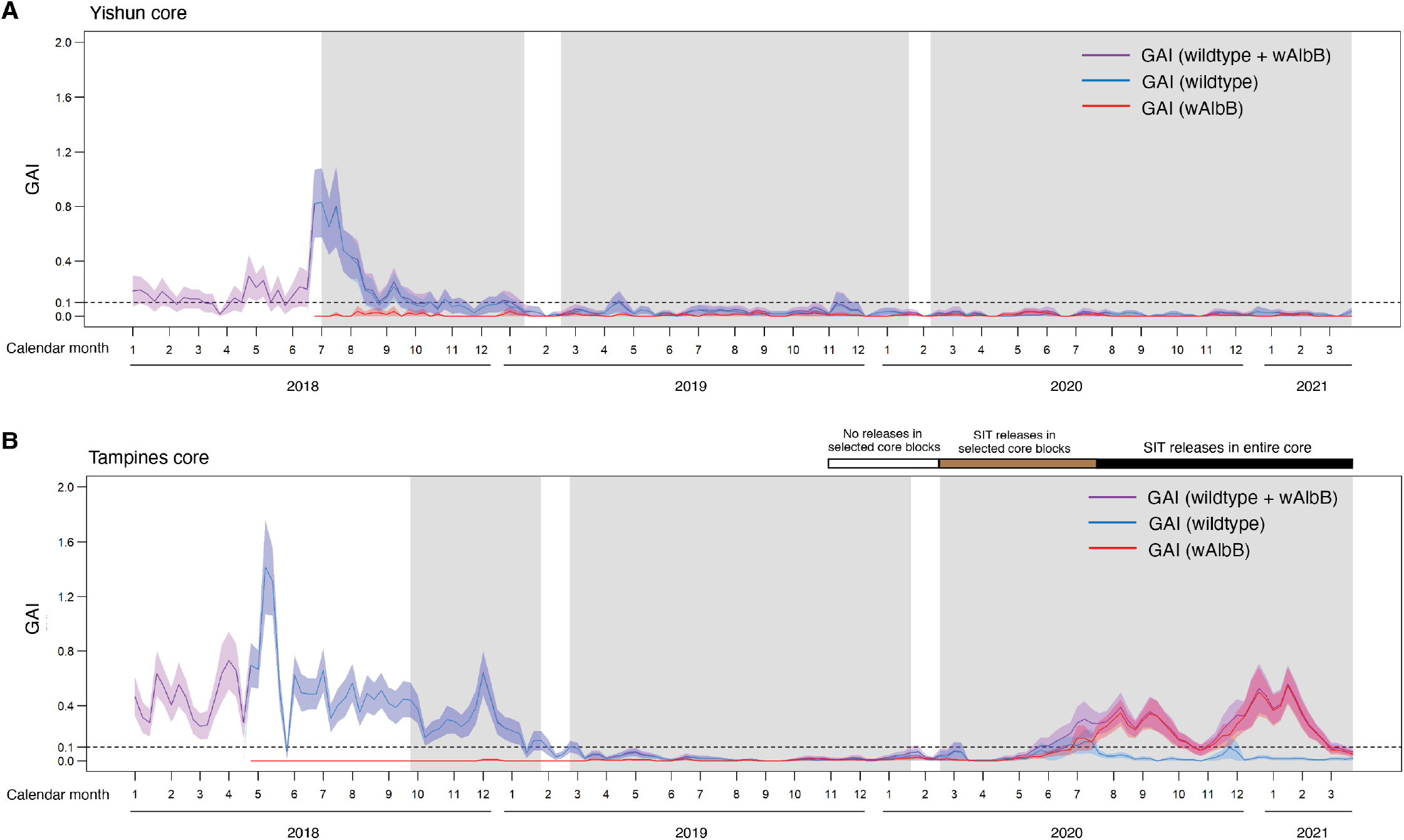
Prevention and mitigation of *w*AlbB establishment. Weekly total, wildtype, and *w*AlbB GAI (average number of female *Aedes aegypti* caught per Gravitrap in the (**A**) Yishun and (**B**) Tampines cores from Jan 2018 to Mar 2021. 95% CIs are shown. Grey-shaded regions indicate release periods.

In Tampines, where pipeline-sorted *w*AlbB-SG males were released, we detected *w*AlbB-positive adult females sporadically and at lower frequencies than in Yishun (Fig. 6B). From May 2020 onwards, however, following a year of very low wildtype *Aedes aegypti* population, we observed a steady increase in *w*AlbB-positive adult females caught (Fig. 6B) in the Tampines core, along with a rebound in the wildtype *Aedes aegypti* population in the same area. This suggests that even with high-fidelity sorting, inadvertent release of a few fertile females can lead to stable establishment of *Wolbachia* in the field, given the lack of competition from the nearly eliminated wildtype population. While we cannot completely rule out the possibility that establishment stemmed from fertile *w*AlbB-SG females released during Phase 1, prior to the use of high-fidelity sex sorting, we consider this unlikely due to the long period (38 weeks) of no detection of *w*AlbB-SG females following the end of Phase 1, as well as the 16-month interval between Phases 1 and 2.

*w*AlbB establishment has no adverse public health impact, since *Wolbachia-*infected mosquito populations are less able to transmit dengue and other *Aedes-*borne pathogens (*9, 10, 36*). Nevertheless, the establishment hampered our use of the *w*AlbB-SG strain for Wolbachia*-*mediated IIT in Tampines to suppress *Aedes aegypti*. We therefore mitigated the establishment via SIT, implementing an X-ray irradiation protocol that sterilizes *w*AlbB-SG males (∼94% male sterility) prior to high-fidelity sorting and release. *w*AlbB-SG males irradiated for male sterility showed reduced fitness compared to *w*AlbB-SG males irradiated for sterilization of residual females (i.e. the protocol used for IIT-SIT in Yishun) (Fig. S1J), as male sterility requires a higher effective dose of irradiation.

Release of sterile *w*AlbB-SG males suppressed both the wildtype and *w*AlbB-positive GAI in the Tampines core to <0.1 (Fig. 6B). Suppression was achieved with the release of sterile *w*AlbB-SG males throughout the entire Tampines core (implemented from July 2020 onwards). The longer time taken to suppress the *w*AlbB-infected *Aedes aegypti* population, as compared to the time taken by IIT-SIT, may be attributed to the fitness cost of SIT irradiation (Fig. S1J) and the incomplete male sterility induced by SIT irradiation (in contrast with the 100% CI conferred by IIT). Rainy monsoon weather towards the end of 2020 may also have created opportunistic breeding habitats, leading to the rebound seen in November. Earlier pausing of releases in the core (from Nov 2019 to Feb 2020), and targeted SIT releases in selected apartment blocks with higher frequencies of *w*AlbB-positive females (from February to July 2020), did not arrest the rise in *w*AlbB-positive females (Fig. 6B), although this lack of effect could also have been due to the relatively low numbers of released males as well as other environmental factors.

As expected with a *w*AlbB-SG population, dengue incidence in the Tampines core remained low in 2020 compared to control sites (incidence rate ratio 0.49, 95% CI 0.32-0.72, p<0.001). All nine dengue-infected *w*AlbB-SG females caught in the Tampines core had very low viral loads of 2.0 × 10^2^ – 1.2 × 10^4^ virus copies per mosquito. On average, this was six logs lower than the viral load of the one infected wildtype *Aedes aegypti* female caught in the same area (5.68×10^9^ virus copies per mosquito).

## Discussion

We demonstrated that *Wolbachia-*mediated IIT can achieve high (>90%), sustained suppression of two wild populations of *Aedes aegypti* and a corresponding reduction in dengue incidence in high-rise public housing estates in Singapore. The highly urbanized, densely populated, and high-rise nature of our trial sites pose unique challenges that are distinct from other pilot trials (*11*–*13*). Lessons learned in our study will therefore be applicable to cities looking to use IIT to reduce dengue.

Both *Wolbachia*-mediated IIT strategies used in the study—IIT-SIT in Yishun and high-fidelity sex-sorting (*13*) in Tampines—resulted in a profound suppression of mosquito populations. Release of IIT-SIT *w*AlbB-SG males in Yishun (at release ratios comparable to or lower than pipeline-sorted *w*AlbB-SG males in Tampines) was effective, although with a longer lead time to suppression due to the fitness cost associated with irradiation. Singapore’s largely successful vector control programme (*21*) has kept the *Aedes aegypti* population relatively low compared to other tropical cities, with an *Aedes* house index typically <1% (*23*). Our data suggest that in this context, relatively less fit IIT-SIT males released in sufficient numbers and according to an effective release plan (e.g. ensuring good spatial and temporal distribution; releasing early in the morning during peak mosquito activity) can compete successfully for urban females. However, reduced fitness is likely to pose a challenge in settings with much higher starting target populations and/or restrictions on the number of released males. Programs in such settings could consider first releasing non-irradiated IIT males to plunge initially very high populations. However, as vector numbers fall close to elimination and the likelihood of stable establishment concurrently rises, additional measures are needed to ensure no establishment of *Wolbachia* in the field, so that *Wolbachia*-mediated IIT continues to be effective for population suppression.

We found that expansion of the release area to create or enlarge buffer zones around the cores was associated with sustained maintenance of >90% suppression in the cores (Fig. 2, 3), and allowed sustained low populations in both sites with no or reduced numbers of *w*AlbB-SG males released, suggesting that suppression can be more effective with larger release sites. This is supported by our compartmental model, which suggests that immigration of wildtype *Aedes aegypti* from contiguous non-release sites plays a role in preventing elimination of *Aedes aegypti* in release sites within the current geographical area of study. As IIT programs scale up, the ability to maintain suppression over large areas with smaller release numbers is expected to reduce costs, improve sustainability, and minimize inconvenience to residents; avoiding excess release numbers should also reduce the likelihood of *Wolbachia* establishment due to the smaller number of females inadvertently released (*33*). With an effective monitoring system such as a network of traps (*34, 37*), programs could transition from regular releases to a risk-based approach, where releases are only performed when and where the vector population exceeds a certain threshold, or where releases are adapted to mosquito trap indices in an operationally manageable manner (*33*).

The detection of a positive spillover impact on dengue incidence and *Aedes aegypti* population in non-release areas up to 1km from the cores (Fig. 4, S5) demonstrated an added advantage of IIT. The spillover could be explained by the flight range of *w*AlbB-SG males (we recaptured *w*AlbB-SG males up to 230m away from release points, and genetic analysis (*38*) suggests that wildtype local mosquitoes may disperse as far as 400-500m), and movement of people. Reducing dengue transmission in the typically high-risk release zones may have reduced the likelihood of viral introduction into adjacent zones through infected people moving across zones. Further analysis of human movement between release and non-release areas is needed to assess this hypothesis. Likewise, dengue cases reported from release sites could have acquired the disease from adjacent sites or elsewhere, and the reduction in dengue cases is thus expected to be enhanced as the release site expands. As a large number of dengue infections are clinically inapparent, dengue incidence data alone do not capture all infections averted due to the *Wolbachia* intervention. Longitudinal cohort studies examining seroconversion may allow us to better assess the full epidemiological impact of IIT (*39*).

Our data further show that when the wildtype mosquito population is suppressed to very low levels—possibly close to elimination, as in the Tampines core—release of even a few fertile *w*AlbB-SG females could result in establishment of *w*AlbB in the field population. This threshold may be as low as three individuals, the minimum number of *w*AlbB-SG females we believe were released in the Tampines core during Phase 2. In Yishun, the use of irradiation to sterilize residual females was effective at preventing establishment of *w*AlbB and a subsequent surge in the *w*AlbB-SG population. During a suppression program, unintentional release of a few fertile females is likely regardless of the methodology used, and thus having an alternative approach for preventing wAlbB establishment, especially when wildtype populations are low, or to mitigate wAlbB establishment is essential.

If establishment does occur, our study demonstrated that an SIT approach can be used to mitigate it, albeit with a longer lead time due to the reduced fitness of X-ray-sterilized *w*AlbB-SG males and their imperfect sterility (as compared to 100% when based on CI). Alternatively, release of a second bi-directionally incompatible strain which also exhibits CI with wildtype mosquitoes as well as with the established Wolbachia release strain and preserves the high competitiveness of non-irradiated males could be considered to suppress the *w*AlbB-established population (*40*).

Further study is needed to determine if SIT or a bi-directionally incompatible strain can eradicate the establishment in the Tampines core. However, the area currently appears to benefit from a *w*AlbB-infected *Aedes aegypti* field population that is both dengue resistant and well suppressed. This “hybrid” approach aligns with the Singapore national vector control program’s focus on reducing mosquito populations (*21*); achieves the same outcomes as an IIT strategy, i.e. dengue reduction coupled with reduced biting pressure; and, due to the low mosquito population, likely reduces the risk of accumulation of dengue virus variants able to escape *Wolbachia-*mediated blocking. Such a hybrid approach could be explored for areas with a very high dengue burden, which may benefit from initial releases of non-irradiated IIT males to quickly plunge high vector populations; any resulting *Wolbachia-*infected, dengue-resistant population could then be suppressed with SIT or a bi-directionally incompatible strain.

The average medical and indirect cost of dengue in Singapore has been estimated at about US$50 million per year during the decade of 2000-2009 (*41*), which saw an average annual reported dengue case count of 5,893. This cost would have increased in the most recent decade (2011-2020), during which an annual average of 13,220 cases were reported; the dengue burden and associated costs are expected to rise further in the future as Singapore’s urban areas expand to accommodate an increasing population. A detailed cost-effectiveness analysis of our IIT program will be conducted when robust data on the required level of geographical coverage, epidemiological impact, and future costs of mass-rearing and release are determined. Economies of scale and the incorporation of automation or other novel technologies into the mass-rearing and release processes are likely to improve long-term cost-effectiveness.

Even as IIT programs scale up for future deployment, IIT should not be viewed as a complete replacement for conventional vector control methods. Public health authorities would do well to continue to set aside sufficient resources and capacity for continued source reduction efforts, including community engagement to maintain high public vigilance against mosquito breeding and prevent complacency. In our experience, both the entomological and epidemiological impacts of IIT are likely to be maximized if IIT is used to complement and enhance, rather than to replace, conventional vector control measures.

## Data Availability

All data is available in the main text or the supplementary materials.

## Acknowledgments

We thank Tim Barkham, Christl Donnelly, Neil Ferguson, Duane Gubler, Ary Hoffmann, and Vernon Lee (Dengue Expert Advisory Panel, National Environment Agency, Singapore) for advice on the study design and manuscript; and Jérémy Bouyer and Andrew Parker (International Atomic Energy Agency) for technical guidance on radiation-based SIT.

## Author contributions

Conceptualization: LCN, CHT, CSC. Methodology: LCN, CHT, CSC, YN, DL, TC, AC, LYT, CL, JWPS, ILM, HY, SYK, MFZ, BJW, EGK, NS, LU, DDY, ARC, BD, SH. Software: YN, LYT, FL, JO, AS, BD, SH. Validation: CHT, YN, LYT, FL, BH. Formal analysis: CHT, CSC, YN, SS, AC, LYT, SYK, JO, AS. Investigation: CHT, CSC, YN, DL, AY, TC, AC, LYT, CL, JWPS, ILM, HY, SYK, MFZ, FL, KV, LTS, CLW, BJW, EGK, YY, BH, NS, LU. Resources: JWPS, ILM, BJW, EGK, YY, NS, LU, DDY, ZX. Data curation: YN, LYT, JO, AS, EGK, YY, Writing – original draft: LCN, CHT, SS. Writing – review & editing: LCN, CHT, CSC, SS, BJW, EGK, ARC, BD, SH. Visualization: YN, SS, LYT, JO, AS, LTS,. Supervision: LCN, CHT, CSC, YN, ARC. Project administration: LCN, CHT, CSC, SS, KV, LTS, CLW, BJW, EGK, YY. Funding acquisition: LCN, CHT, CSC, SS.

## Data and materials availability

All data is available in the main text or the supplementary materials.

## Supplementary figures and tables

**Figure S1.**
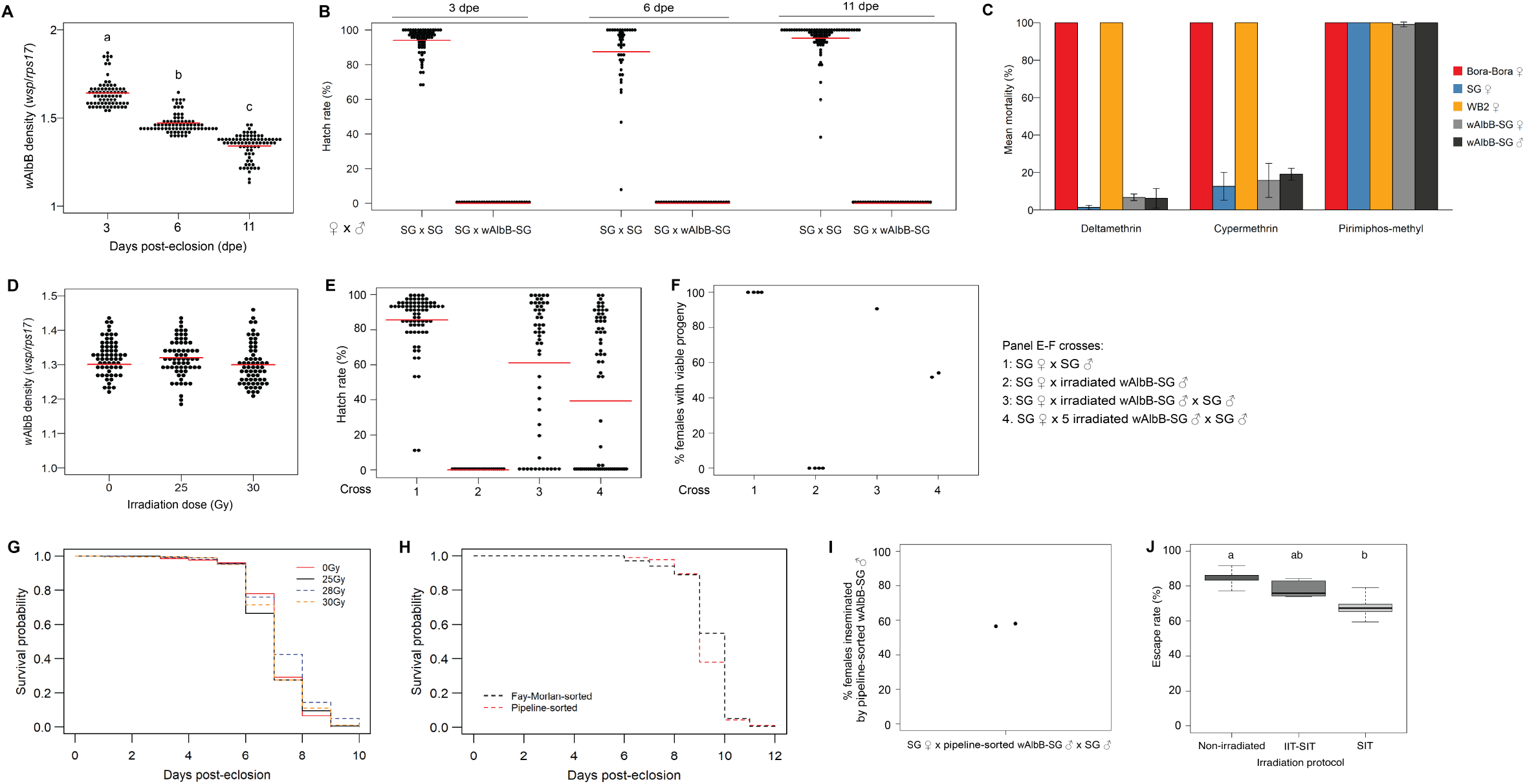
Characterization of the *w*AlbB-SG *Aedes aegypti* line. **(A)** *w*AlbB density in *w*AlbB-SG male adults at varying days post-eclosion. Each data point represents an individual *w*AlbB-SG male; red horizontal bars indicate means. Groups denoted by different letters are significantly different from each other (p<0.05 by Kruskal-Wallis and Dunn’s post-hoc tests). **(B)** Hatch rates from crosses between *w*AlbB-SG and SG lines at varying days post-eclosion of males (n=3 for each cross; bars indicate means and error bars indicate SEM). **(C)** Mean mortality of *w*AlbB-SG, its parental strain WB2, the wildtype SG strain, and the insecticide-susceptible Bora-Bora strain upon exposure to insecticides via the WHO standard bioassay. For each insecticide, three tests with four replicates each were performed on three consecutive days. **(D)** *w*AlbB density in non-irradiated (0 Gy) and irradiated *w*AlbB-SG males. Each data point represents an individual *w*AlbB-SG male; red horizontal bars indicate means. **(E-F)** Hatch rates of eggs laid by SG females **(E)** and proportion of SG females producing viable progeny **(F)** following competitive mating with irradiated (30 Gy) *w*AlbB-SG males and non-irradiated SG males at 1:1 and 5:1 ratios (crosses 3 and 4). Control compatible (SG males x SG females, cross 1) and incompatible (irradiated *w*AlbB-SG males x SG females, cross 2) crosses are also shown. In panel (F), each data point represents a biological replicate. **(G-H)** Adult survival probability curves of *w*AlbB-SG males exposed to varying doses of X-ray irradiation at the pupal stage **(G)** and of Fay-Morlan- and pipeline-sorted *w*AlbB-SG males **(H). (I)** Proportion of SG females inseminated by pipeline-sorted *w*AlbB-SG males following competitive mating with pipeline-sorted *w*AlbB-SG males and Fay-Morlan-sorted SG males at a 1:1 ratio. **(J)** Flight test escape rates of *w*AlbB-SG males subjected to different irradiation protocols. For each treatment condition, two rounds of flight assays with three replicates each were carried out (n=6). Lines and limits of the boxes indicate medians and 25^th^ and 75^th^ percentiles respectively; whiskers extend to 1.5 times the interquartile range. Groups denoted by different letters are significantly different from each other (p<0.05 by Kruskal Wallis and Dunn’s post-hoc tests).

**Figure S2.**
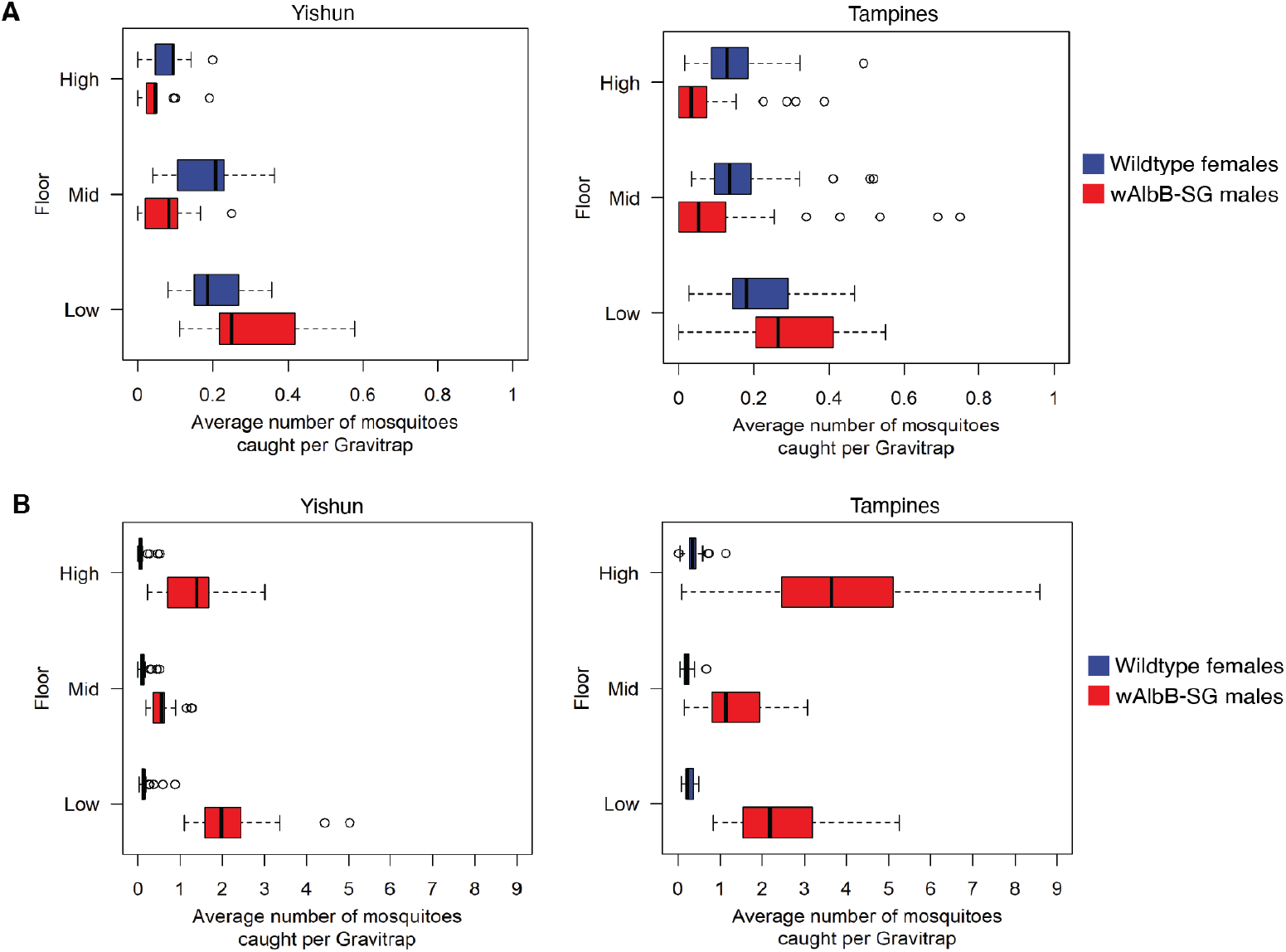
Vertical distribution of released *w*AlbB-SG males. Vertical distribution of *w*AlbB-SG males and wildtype *Aedes aegypti* females among low (levels 1-4), mid (levels 5-8), and high (levels 9 and above) floors of high-rise apartment blocks after ground-only releases **(A)** and ground and high-floor releases **(B)**. Lines and limits of the boxes indicate medians and 25^th^ and 75^th^ percentiles respectively; whiskers extend to 1.5 times the interquartile range. Outliers are shown as points.

**Figure S3.**
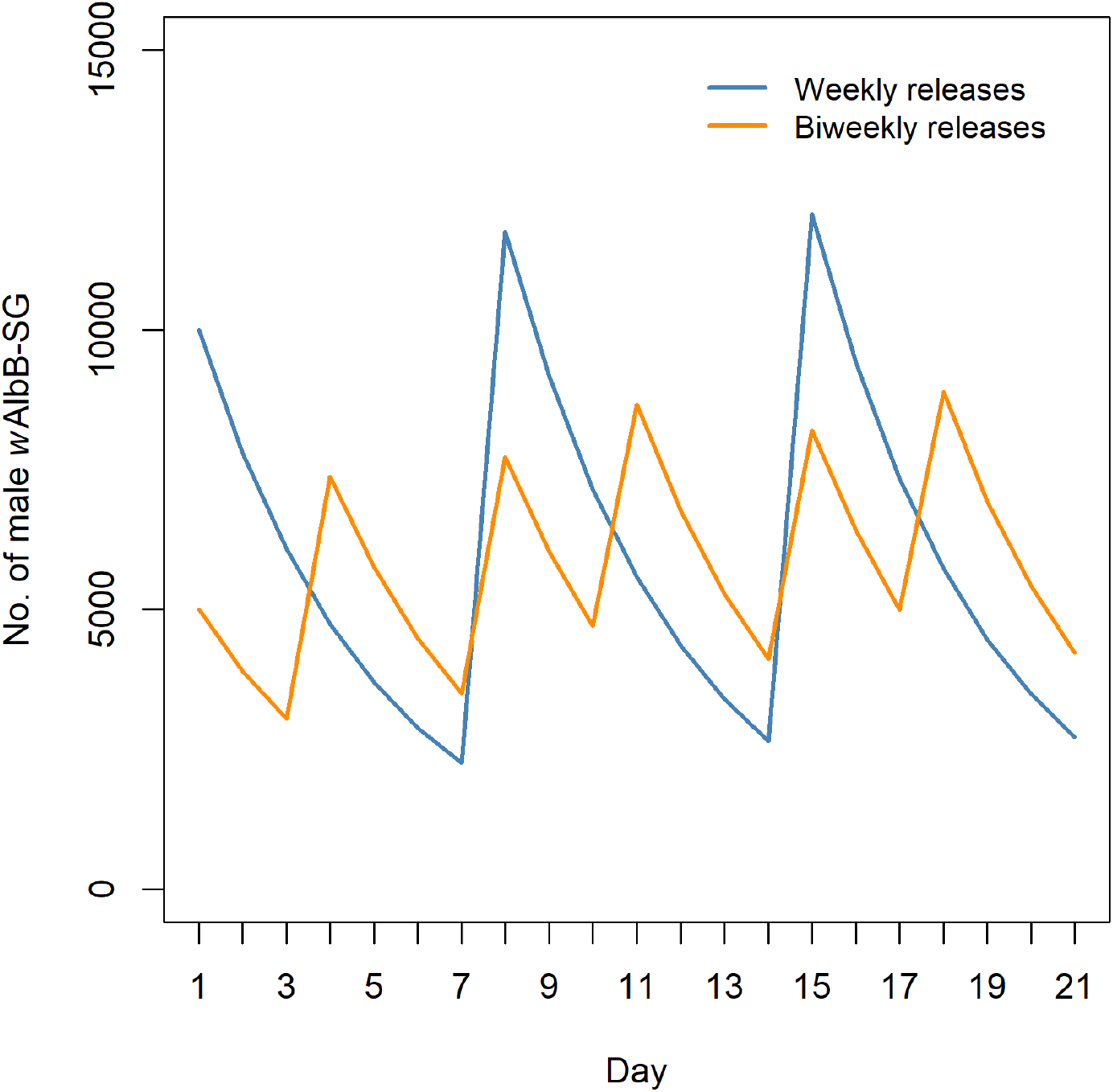
Estimated daily field populations of *w*AlbB-SG males under weekly and biweekly release scenarios. Daily field populations were estimated using a probability of daily survival of 0.78 and a release number of 10,000 *w*AlbB-SG males per week.

**Figure S4:**
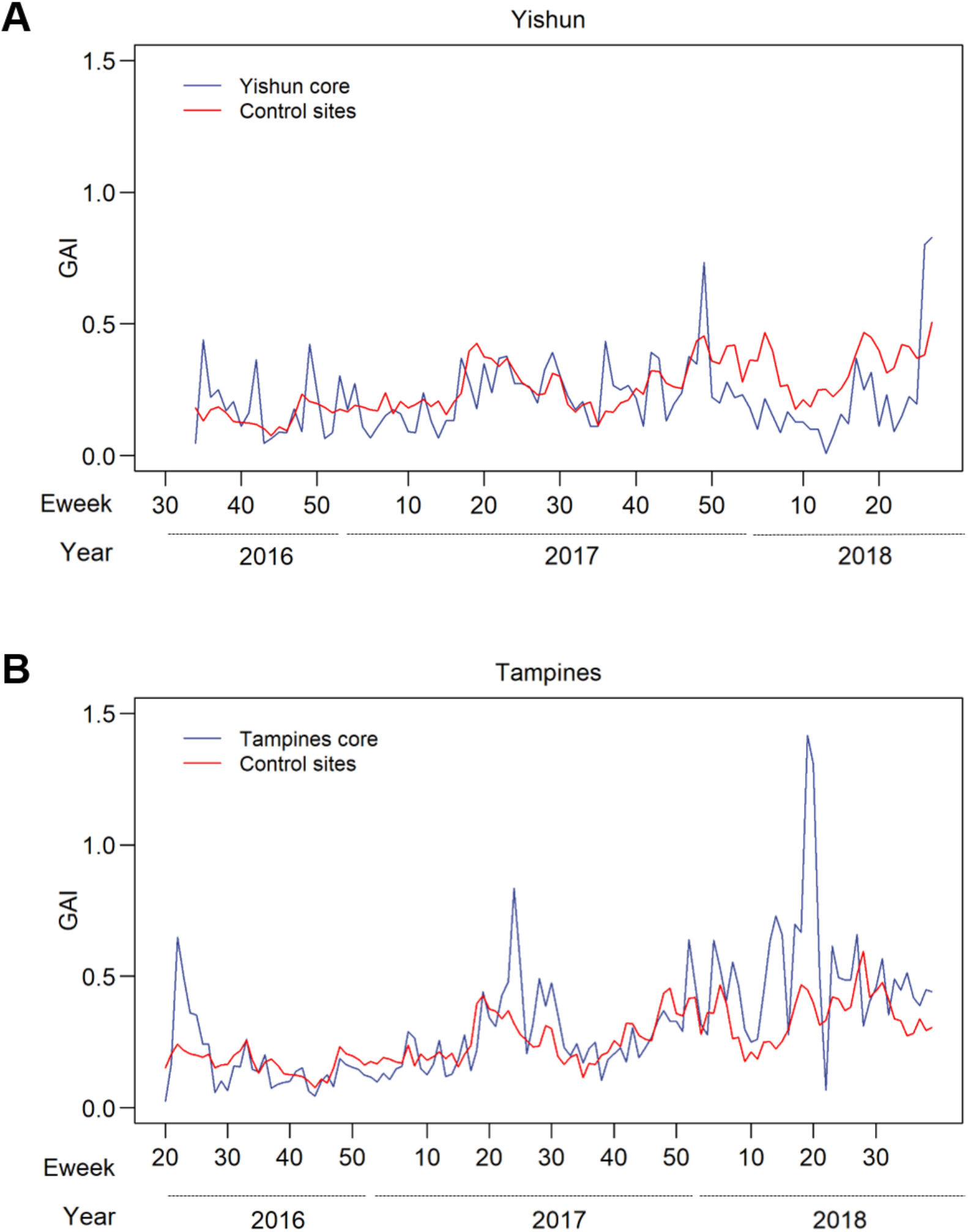
Correlation of baseline (pre-release) GAI in control and **(A)** Yishun and **(B)** Tampines core sites. Pearson’s R for Yishun and Tampines was 0.44 (p<0.05) and 0.63 (p<0.05) respectively.

**Figure S5.**
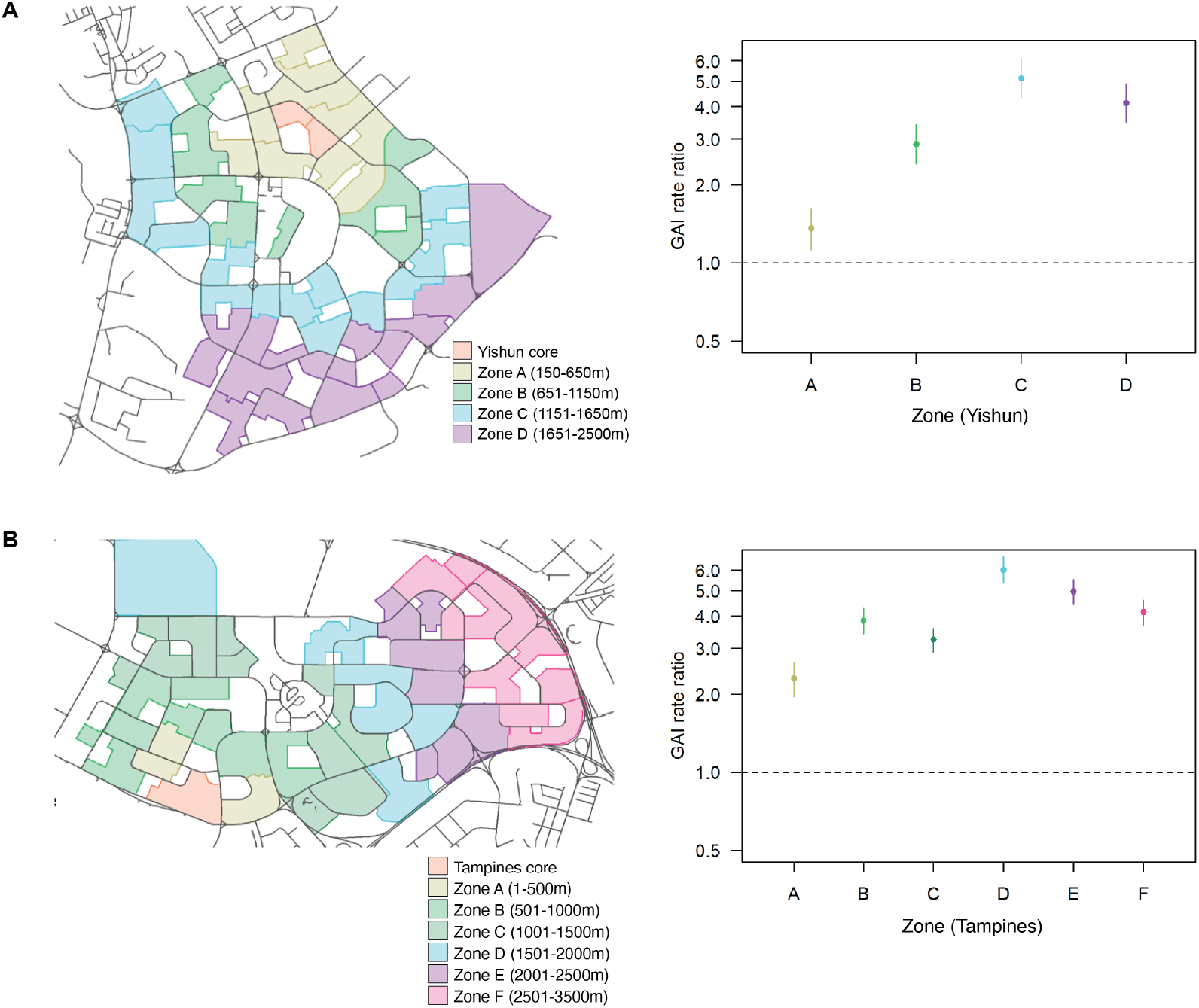
Spillover effect of releases of *w*AlbB-SG males. **(A-B)** Rate ratios of GAI in non-release zones relative to core release zones in Yishun **(A)** and Tampines **(B)**. Point estimates and 95% confidence intervals are shown.

**Figure S6:**
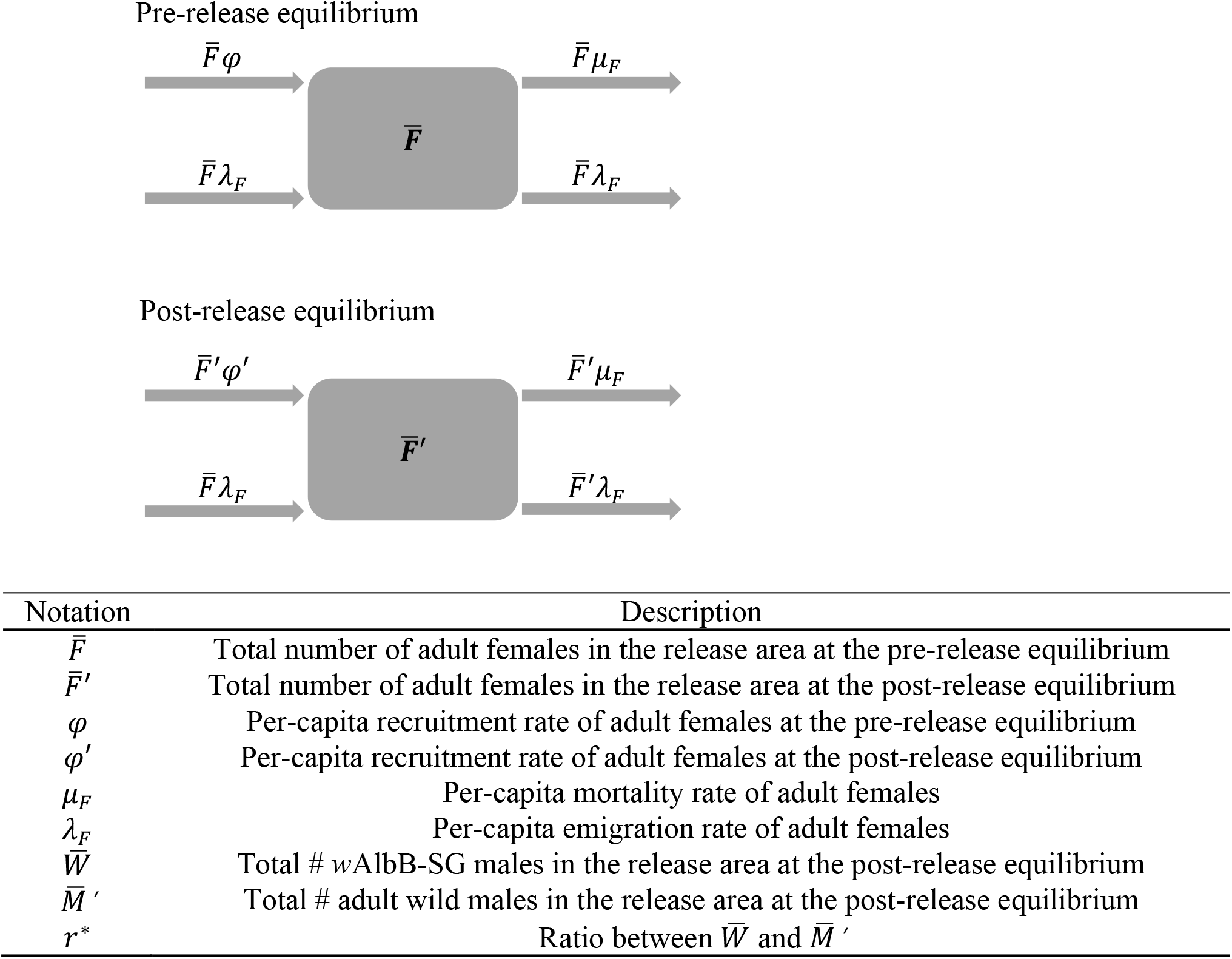
Visualization and parameters of compartmental model developed to estimate immigration size. Compartments 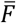 and 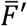 denote the total number of adult females in the release area at the pre- and post-release equilibriums respectively. Each adult female in the release area had a daily mortality rate of *μ*_*F*_ and a daily emigration rate of *λ*_*F*_, both assumed to be constant over time. We used the term “total recruitment rate” to refer to the total rate at which female pupae in the release area became adults. This was assumed to be proportional to the total number of adult females in the release area with the proportionality coefficients *φ* and *φ*′ denoting the per-capita recruitment rate of adult females at the pre- and post-release equilibriums respectively. For the pre-release equilibrium the following pairs of parameters were assumed to be equal: the total recruitment rate 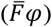 and mortality rate 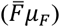, and the total immigration rate and emigration rate 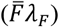. We also assumed the total immigration rate to be constant over time despite the substantial decrease in the total emigration rate owing to the suppression effect. At the post-release equilibrium, the per-capita recruitment rate *φ*′ was given by

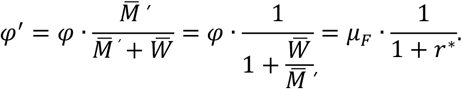

The parameters 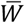 and 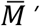 denote the total numbers of *w*AlbB-SG and adult wild males present in the release area at the post-release equilibrium, and their ratio is denoted by *r*^∗^. We assume near-equal mating competitiveness between the wild and *w*AlbB-SG males together with perfect cytoplasmic incompatibility, and the per-capita recruitment rate at the post-release equilibrium becomes 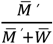 times as large as the pre-release rate *φ*. As we assumed that the recruitment and mortality rates were equal prior to the release, we replace *φ* with the daily mortality rate of female *Aedes aegypti* (*μ*_*F*_), based on Brady et al.’s estimate (*42*). Despite the total recruitment rate being lower than the total mortality rate post-release, the wild population in the release area did not die out completely due to net immigration. At the post-release equilibrium,

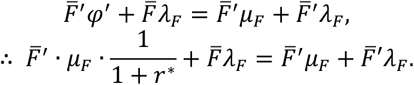

Therefore, the daily per-capita emigration rate *λ*_*F*_ Is

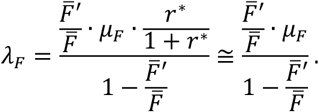

The above approximation assumed that the ratio 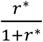 was very close to one. The denominator was approximated using the overall suppression effect estimate at the end of the intervention period for the whole release area at each site.

**Figure S7:**
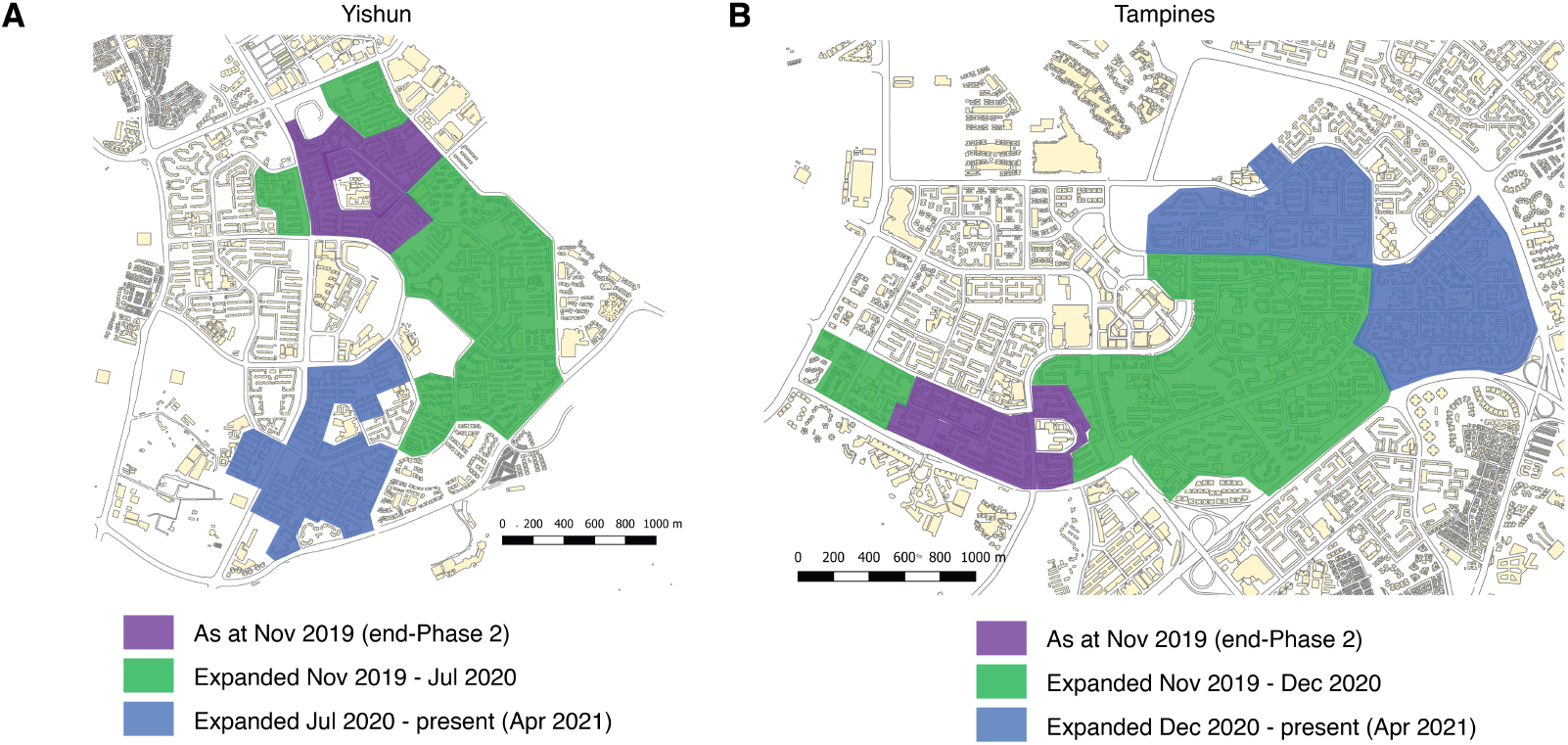
Expansion of release sites. Colored areas indicate release areas.

**Table S1:** Sensitivity analysis of BACI suppression estimates in the Yishun and Tampines cores. BACI suppression estimates in the Yishun and Tampines cores for the final month of Phase 2 are shown, relative to all 7 control sites or each control site individually. * indicates statistical significance (p<0.05).

| **Control site (referent)** | **Pairwise BACI suppression estimate** |  |
| --- | --- | --- |
|  | **Yishun core** | **Tampines core** |
| All 7 control sites | -92% | -98%* |
| Bishan St 11 | -93%* | -98%* |
| Commonwealth Close | -89% | -98%* |
| Jurong East St 21 | -91% | -95%* |
| Marine Terrace | -95%* | -98%* |
| Serangoon Central | -79% | -97%* |
| Tampines St 23 | -87% | -94%* |
| Toa Payoh | -95%* | -98%* |

**Table S2.**
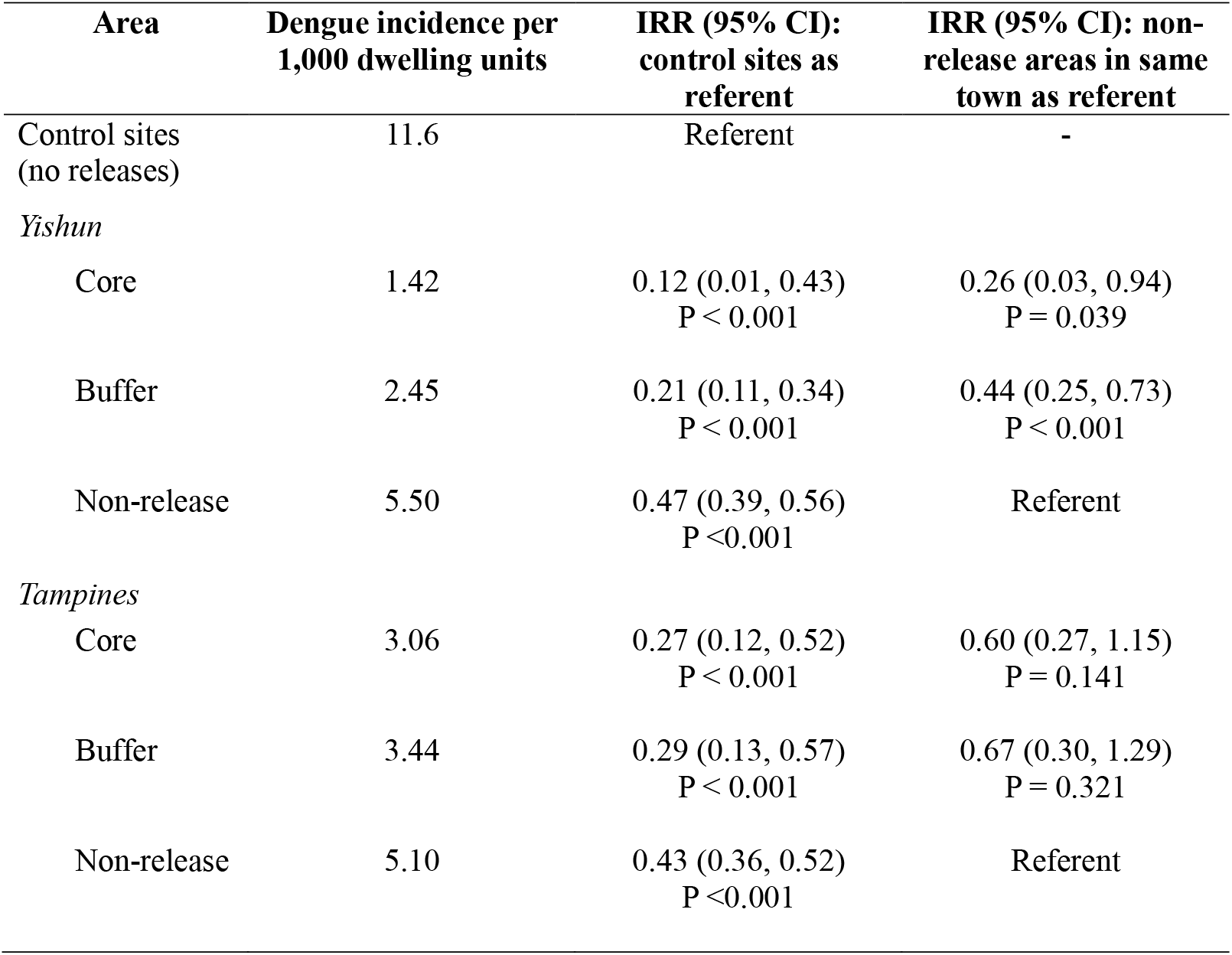
Reduction in 2019 dengue incidence at intervention sites. Incidence rate ratios (IRRs) were computed relative to either the control sites or to non-release areas within the same town.

| Area | Dengue incidence per 1,000 dwelling units | IRR (95% CI): control sites as referent | IRR (95% CI): non-release areas in same town as referent |
| --- | --- | --- | --- |
| Control sites (no releases) | 11.6 | Referent | - |
| *Yishun* |  |  |  |
| Core | 1.42 | 0.12 (0.01, 0.43)<br>P < 0.001 | 0.26 (0.03, 0.94)<br>P = 0.039 |
| Buffer | 2.45 | 0.21 (0.11, 0.34)<br>P < 0.001 | 0.44 (0.25, 0.73)<br>P < 0.001 |
| Non-release | 5.50 | 0.47 (0.39, 0.56)<br>P < 0.001 | Referent |
| *Tampines* |  |  |  |
| Core | 3.06 | 0.27 (0.12, 0.52)<br>P < 0.001 | 0.60 (0.27, 1.15)<br>P = 0.141 |
| Buffer | 3.44 | 0.29 (0.13, 0.57)<br>P < 0.001 | 0.67 (0.30, 1.29)<br>P = 0.321 |
| Non-release | 5.10 | 0.43 (0.36, 0.52)<br>P < 0.001 | Referent |

**Table S3.**
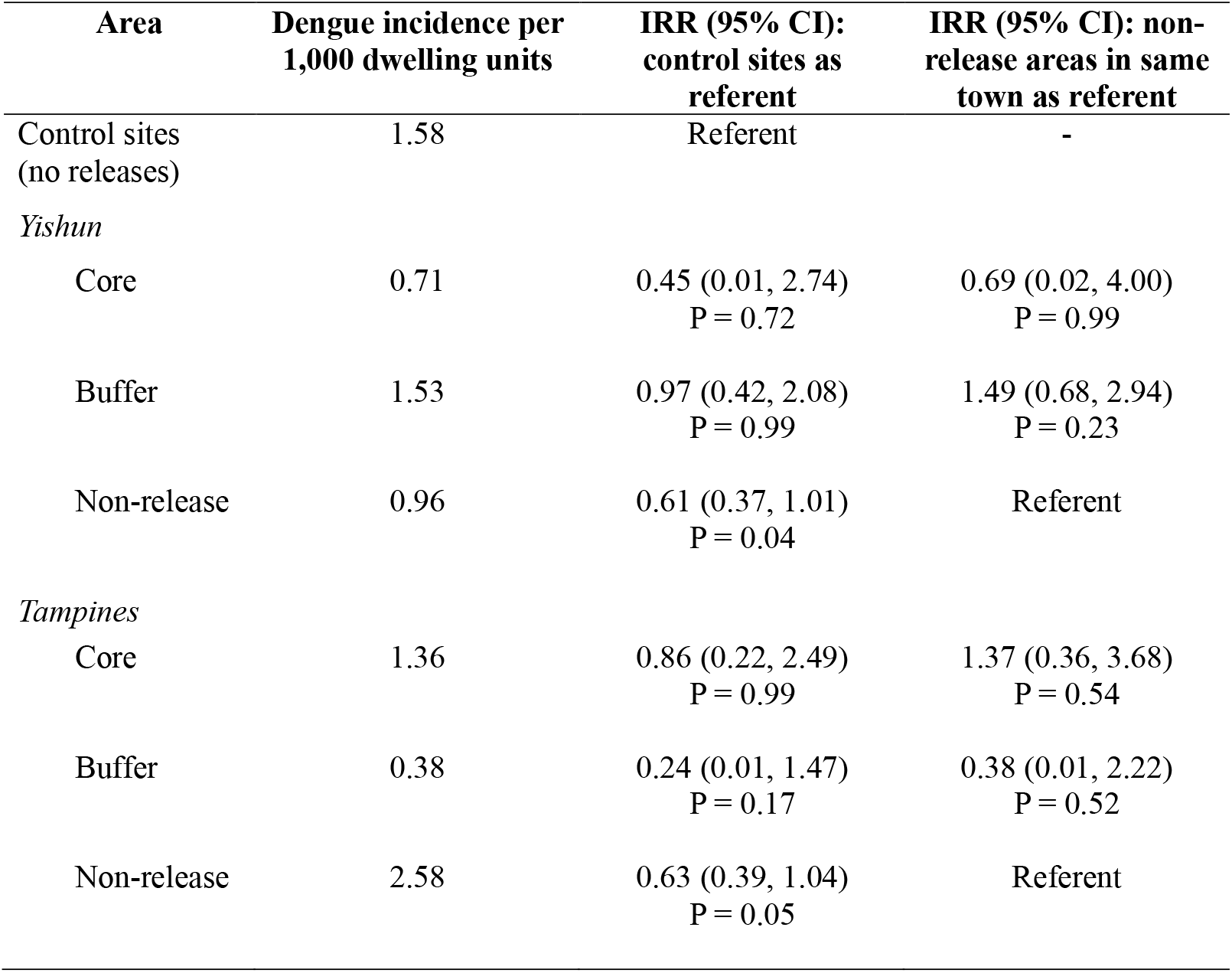
Pre-trial (2017) dengue incidence at intervention sites. Incidence rate ratios (IRRs) were computed relative to either the control sites or to non-release areas within the same town.

| Area | Dengue incidence per<br>1,000 dwelling units | IRR (95% CI):<br>control sites as<br>referent | IRR (95% CI): non-release areas in same<br>town as referent |
| --- | --- | --- | --- |
| Control sites<br>(no releases) | 1.58 | Referent | - |
| *Yishun* |  |  |  |
| Core | 0.71 | 0.45 (0.01, 2.74)<br>P = 0.72 | 0.69 (0.02, 4.00)<br>P = 0.99 |
| Buffer | 1.53 | 0.97 (0.42, 2.08)<br>P = 0.99 | 1.49 (0.68, 2.94)<br>P = 0.23 |
| Non-release | 0.96 | 0.61 (0.37, 1.01)<br>P = 0.04 | Referent |
| *Tampines* |  |  |  |
| Core | 1.36 | 0.86 (0.22, 2.49)<br>P = 0.99 | 1.37 (0.36, 3.68)<br>P = 0.54 |
| Buffer | 0.38 | 0.24 (0.01, 1.47)<br>P = 0.17 | 0.38 (0.01, 2.22)<br>P = 0.52 |
| Non-release | 2.58 | 0.63 (0.39, 1.04)<br>P = 0.05 | Referent |

## Supplementary Materials

### Materials and Methods

#### Generation of *w*AlbB-SG line

To generate *w*AlbB-SG, a localized *Aedes aegypti* line stably infected with *Wolbachia*, we outcrossed 500 females from the WB2 line (a *w*AlbB-infected *Aedes aegypti* line) with an equal number of F3 males from the SG line (a wildtype *Aedes aegypti* line derived from field collections in Singapore). Resultant hybrid females were outcrossed with an equal number of F3 SG males; this process was repeated a total of six times to obtain *w*AlbB-SG. Mosquito lines were maintained under standard insectary conditions at 27±1°C and 75±10% relative humidity (RH), with a 12h:12h light:dark cycle. For colony maintenance and experimental procedures, females were fed on commercially obtained specific pathogen-free pig’s blood using a Hemotek device (Discovery Workshops, UK).

#### Cytoplasmic incompatibility (CI)

To confirm CI, 50 *w*AlbB-SG males and 50 SG females were allowed to mate for 24 hours inside a 30 x 30 x 30 cm cage. Control cages of 50 SG males and 50 SG females were set up under similar conditions. Three replicates each of experimental and control conditions were performed. Females were then bloodfed as described above and transferred individually to ovipots containing germination paper. After 48 hours, 2 ml deionized water was added to wet the ovipot and allow mosquitoes to oviposit. After another 72 hours, germination papers containing eggs were collected and incubated at 28±1°C and 80±10% RH for 3 days to allow embryogenesis. Germination papers were then submerged in hatching broth (0.071g baker’s yeast (MP Biomedicals, France), 0.357 g nutrient media (CM0001 Nutrient broth, Oxoid, England) and 1000 ml deionised water) to induce hatching. After 24 hours, germination papers were removed and allowed to dry for another 72 hours before re-submerging to allow any remaining unhatched eggs to hatch. To determine hatch rate, L2 or L3 larvae were counted 72 hours post-hatching. Spermathecae of female mosquitoes that laid eggs that did not hatch were dissected to confirm insemination. Data from female mosquitoes without any observable sperm in their spermathecae were excluded from the analysis. To assess the impact of age on CI, the assay was performed with 3-, 6-, and 11-day-old *w*AlbB-SG males.

#### *w*AlbB maternal transmission

To determine maternal transmission efficiency of *w*AlbB, a total of 1,648 adult offspring from 103 *w*AlbB-SG females were assayed for *w*AlbB via qPCR as described below. At least 34 *w*AlbB-SG females from the first-, third, and fifth-generation outcrosses contributed offspring, and at least 16 offspring from each *w*AlbB-SG female were sampled.

#### *w*AlbB detection and quantification

Individual mosquitoes were homogenized in a MM301 mixer mill (Retsch, Germany) and total DNA was extracted using the Qiagen Blood and Tissue kit (Qiagen, Germany). Detection and quantification of *w*AlbB was performed using a multiplex real-time qPCR assay targeting the *w*AlbB *wsp* gene (forward primer 5’ AGYTATGATGTAACYCCAGA-3’, reverse primer 5’-TTAAACGCTACTCCAGCTTCT-3’, probe FAM-GGTTCTTATGGTGCTAGTTTTAATAAAGA-BHQ1) and the *Aedes aegypti* RpS17 gene (forward primer 5’-TCCGTGGTATCTCCATCAAGCT-3’, reverse primer 5’-CACTTCCGGCACGTAGTTGTC-3’, probe TR-CAGGAGGAGGAACGTGAGCGCAG-BHQ2). qPCR reactions were performed in 20ul total volume containing 1X SensiFAST No-Rox Probe Mastermix (Bioline, USA); 0.4uM and 0.2uM *wsp* primers and probes, respectively; 0.2uM and 0.1uM RpS17 primers and probes, respectively; and 5ul of DNA template. Cycling was carried out on a Rotorgene Q (Qiagen, Germany) using the following conditions: 95°C for 5 min, 45 cycles at 95°C for 10s, and 60°C for 30 sec. Amplification of the *wsp* and RpS17 genes from individual mosquitoes was compared against a standard curve generated from a 10-fold serial dilution of DNA standard for each gene.

#### Mating competitiveness

To determine mating competitiveness of *w*AlbB-SG males, 10 *w*AlbB-SG males, 10 SG males, and 20 SG females were allowed to mate for 24 hours inside a 30 x 30 x 30 cm cage. The following controls were set up under similar conditions: 20 SG females and 20 SG males (compatible wildtype cross); 20 *w*AlbB-SG females and 20 *w*AlbB-SG males (compatible *w*AlbB-SG cross); and 20 SG females and 20 *w*AlbB-SG males (incompatible cross). Five replicates were performed for the competitiveness trial, and 2 replicates for each of the control crosses. Males were released first into the cages and allowed to acclimatize for 1 hour before females were introduced. After mating, females were bloodfed and allowed to oviposit; the hatch rate of the resulting eggs was then determined as described above. Mating competitiveness of *w*AlbB-SG males was determined by computing the Fried competitiveness index C as follows: C = (w/s) * [(H_w_ -H_c_) / (H_c_ -H_s_)], where w and s refer to the number of SG (wildtype) and *w*AlbB-SG males respectively in each competitive cage; H_w_ refers to the number of SG females laying viable eggs in the compatible wildtype cross; H_c_ refers to the number of SG females laying viable eggs in the competitiveness trial; and H_s_ refers to the number of SG females laying viable eggs in the incompatible cross (*27, 43*). C = 1 indicates equivalent mating performance between male *w*AlbB-SG and SG males. Determination of mating competitiveness of irradiated (30Gy) *w*AlbB-SG males was carried out with similar methodology except that mosquitoes were allowed to mate in a 200(L) x 250(H) x 100(W) cm flight cage. Ratios used were (i) 100 each of *w*AlbB-SG males, SG males, and SG females, and (ii) 500 *w*AlbB-SG males, 100 SG males, and 100 SG females.

To compare the mating competitiveness of Fay-Morlan-sorted and pipeline-sorted *w*AlbB-SG males, we used the fluorescent dye rhodamine B (RhB) to directly measure mating interactions between these two groups of *w*AlbB-SG males and SG females, according to the protocol in (*44*). In brief, each group of males was marked in turn with RhB by feeding in sucrose solution. After competitive mating, the presence of fluorescing (RhB-marked) sperms in the spermathecae of a female were used to determine her mating partner. This method allows comparison of mating competitiveness between two *Wolbachia*-infected lines.

To compare the competitiveness of IIT-SIT-irradiated and SIT-irradiated *w*AlbB-SG males, we assessed flight ability of adult mosquitoes using a flight test device (FTD) as a proxy for mating competitiveness (*45*). Around 100 mosquitoes per experimental condition were transferred into the FTD using a mouth aspirator and given two hours to escape from the inner tubes of the FTD. Escape rates were calculated by dividing the number of adults that escaped the inner tubes by the total number of mosquitoes transferred into the FTD at the start.

#### Longevity assays

To determine the impact of irradiation on longevity under laboratory conditions, male *w*AlbB-SG pupae were irradiated at 0, 25, 28, and 30Gy and allowed to eclose. Fifty adult male *w*AlbB-SG of similar age from each irradiation dose were transferred to 30 x 30 x 30 cm cages, and provided with sugar solution for the first two days and water thereafter. Dead adults were recorded and removed on a daily basis. The same process was followed for all longevity assays.

#### Susceptibility of *w*AlbB-SG to insecticides

Insecticide susceptibilities of *w*AlbB-SG, its parental strain WB2, and the wildtype SG strain were determined by exposing 3-5-day-old males and females to technical-grade pyrethroids (deltamethrin and cypermethrin) and organophosphate (pirimiphos-methyl) insecticides, following the WHO standard bioassay (*46*) with slight modifications. Briefly, a batch of 20-25 mosquitoes was aspirated into a plastic holding tube (12 x 4 cm) lined with clean filter paper and held for 30 min, after which any weak and/or dead mosquitoes were removed and replaced. Mosquitoes were then transferred to a horizontal testing tube lined with an insecticide-treated filter paper and exposed to insecticide (3 hours for pyrethroids and 1 hour for pirimiphos-methyl) (*47*). Mosquitoes were returned to the holding tube for recovery, and mortality at 24 hours post-treatment was recorded. The standard insecticide-susceptible Bora-Bora *Aedes aegypti* strain was used as a control. Diagnostic doses (0.014% for deltamethrin, 0.148% for cypermethrin, and 0.212% for pirimiphos-methyl) were determined using the method described in (*47*).

#### Production of *w*AlbB-SG males for release

Eggs were hatched as described above. After 24 hours, larvae were counted using an automated larvae counter (Orinno Technology Pte Ltd, Singapore) and transferred to rearing trays measuring 53 x 40.5 x 10 cm at a density of one larva per ml of deionized water. Larvae were fed with ground Tetramin Flakes (Tetra, Germany) in either slurry or capsule form. For releases in Yishun, six days post-hatching, male pupae were separated from female pupae using the Fay-Morlan glass plate separation method (Wolbaki, China), and allowed to mature for at least 18 hours prior to irradiation for IIT-SIT releases in Yishun (see below). Pupae were allowed to eclose inside release containers. For releases in Tampines, the automated, multi-step sex separation process described in (13) was followed with pupae collected six-days post hatching.

#### Determination of female-sterilizing dose of X-ray irradiation

To determine the level of female sterility induced by X-ray irradiation, female *w*AlbB-SG pupae were irradiated at varying doses (0, 25, 28, and 30 Gy). At least 50 female pupae from each condition were separately transferred to 30 x 30 x 30 cm cages for eclosion. Twenty-four hours post-eclosion, an equal number of adult SG males were transferred to each cage, and mosquitoes were allowed to mate for at least 48 hours. Following a bloodmeal, blood-fed females were transferred to individual 40ml paper cups and allowed to lay eggs; hatch rates were determined as described above. Induced sterility was calculated using the following formula: 100% - (observed hatch rate/control hatch rate).

#### X-ray irradiation of pupae

Irradiation was performed with either the RS2000 (Phase 2) and RS2400V irradiators (beyond Phase 2) (Rad Source Technologies Inc., USA). For IIT-SIT irradiation (female sterilization), pupae were irradiated in plastic petri dishes with diameter 9.4 cm (for longevity and mating competitiveness experiments) or 15.0 cm (for release), or in plastic containers measuring 6.5 cm in diameter and 7.0 cm in height (for release). For SIT irradiation (male sterilization), pupae were irradiated in petri dishes with diameter 9.4 cm. Petri dishes or containers were placed at the center of the irradiation chamber of the X-ray irradiator. The effective dose was 30 Gy to 35 Gy.

#### Field releases of *w*AlbB-SG males

Releases were conducted twice a week (weekdays between 0630-1030 hrs) at designated public locations in the high-rise housing estates. In Phase 1, releases were limited to ground floors, and were conducted at 6-12 equally spaced release locations per apartment block to facilitate even distribution of mosquitoes. In Phase 2, to ensure a more even vertical distribution of mosquitoes, half the ground release locations were shifted to upper floors, alternating between middle floors (levels 5-6) and high floors (levels 10-11).

#### Monitoring of field mosquito populations

Adult *Aedes aegypti* populations in release and control sites were monitored weekly using an average of six Gravitraps (*31*) per apartment block. Gravitraps were placed in public spaces along corridors and were evenly vertically distributed throughout the block, corresponding to a ratio of approximately one trap for every 20 households. All mosquito samples were sexed and morphologically identified, and all *Aedes aegypti* samples were subjected to PCR for *w*AlbB detection. To assess the mating competitiveness of *w*AlbB-SG males, ovitraps were also deployed for 60 weeks pre-, during, and post-Phase 1 and for 94 weeks pre-, during, and post-Phase 2. In total, up to 253 ovitraps were placed for two days per week in half the apartment blocks in release and control sites (Fig. 1A). Eggs collected were hatched and reared to 3^rd^ or 4^th^ larval instars for morphological identification. All *Aedes aegypti* larvae were subjected to PCR for *w*AlbB detection. Eggs that failed to hatch and dead 1^st^ and 2^nd^ larval instars were individually subjected to PCR for molecular speciation and *w*AlbB detection.

#### Compartmental model and estimation of immigration size

We developed a compartmental model to estimate the daily average number of females that migrated from the non-release area to the release area (Fig. S6), which was expressed as a percentage of the total number of females in the release area at the pre-release equilibrium.

#### Statistical analyses

Statistical analyses were performed in MedCalc for Windows (MedCalc, Belgium), SPSS, or R (R Core Team, Vienna, Austria). Kruskal-Wallis and Tukey’s post-hoc tests were used to compare relative *Wolbachia* density among groups. Pearson’s correlation was used to determine the association in pre-release baseline GAI between release and control sites.

A before-after-control-impact (BACI) statistical design (*48*) was used to assess the impact of male *Wolbachia-*infected *Aedes aegypti* releases. Weekly GAI in the treatment and control sites were computed prior to releases (“pre-release”) and during releases (“release”). The effect of male *Wolbachia*-infected *Aedes* aegypti releases was estimated using the BACI contrast, calculated as the difference (treatment vs. control) between the mean differences (release vs. pre-release):

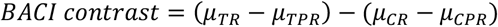

Where µ is the four-week site-specific spatial mean GAI; TR and CR refer to the treatment and control sites during release respectively; and TPR and CPR refer to the treatment and control sites during pre-release respectively.

A controlled interrupted time-series analytical design was used to evaluate the impact of male *Wolbachia*-infected *Aedes aegypti* releases on the *Aedes aegypti* population in the treatment sites. We estimated the change in the average level of GAI following the intervention and the trends of GAI in the pre- and post-intervention period using negative binomial regression. Two separate negative binomial models were fitted for the treatment sites. We included the GAI of the control sites as a covariate in the model to control for secular trends and accounted for seasonality by adding a week variable in the models. In addition, we examined the autocorrelation and partial autocorrelation function plots of the penultimate model before adding lags of the deviance residuals to account for any serial autocorrelation. The general model used to examine the impact of intervention on the *Aedes aegypti* population is as follows:

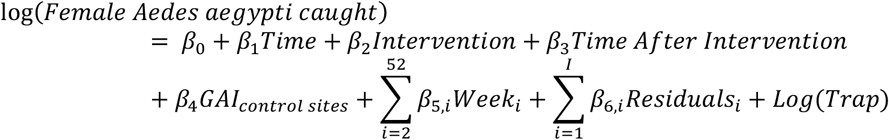

Where *Time* indicates the weekly time interval from the start of the study period, *Intervention* represents the male *Wolbachia*-infected *Aedes aegypti* releases (pre-intervention period coded as 0 and post-intervention period as 1), and *Time After Intervention* indicates the number of weeks after the intervention started (takes on the value zero before the intervention and continuous values following the intervention). *β*_0_ captures the baseline level of the GAI at time *t*=0, *β*_1_ estimates the structural trend in the GAI before intervention (i.e. baseline trend), *β*_2_ estimates the level change in the GAI after the intervention, *β*_3_ reflects the change in trend in the GAI after the intervention. Non-significant variables were removed to achieve parsimony.

To estimate the extent of impact in non-release areas, we computed the Haversine distance from the centroid of the core sector to the centroid of all other non-release sectors for both Tampines and Yishun, and subsequently grouped these non-release sectors into bounded zones using 500m distance intervals (Fig. 4). We computed and compared the dengue incidence and GAI between each zone and the core.

